# Single Cross-Sectional Analysis; Post-Myocardial Infarction Instrumental & Laboratory changes in Prognosis Behavior

**DOI:** 10.1101/2023.02.25.23286453

**Authors:** Basheer Abdullah Marzoog, Natalia Igorevna Bloshkina, Vera Sergeevna Gromova, Ekaterina Vanichkina, Elena Ivanovna Gorshinina

## Abstract

**Background:** Cardiovascular events remain one of the frequently reported causes of death globally.

**Aims:** Assess the early laboratory and instrumental changes in systemic manner in patients with myocardial infarction (MI).

**Objectives:** Patients with fresh myocardial infarction that has been occurred in less than 30 days before hospitalization in the rehabilitation hospital.

**Materials and methods:** A single large-scale retrospective cohort sectional study for patients with fresh myocardial infarction for the period 2014-2019, data collected from Mordovia Rehabilitation Hospital. The study involved 154 patients and analysed 76 parameters for each patient. The t test, Pearson’s correlation coefficient, and ROC test have been used. For statistical analysis used Statistica program.

**Results:** The sample included 154 patients with a history of fresh myocardial infarction. Of 154. 51(33.11%) female and 103 (66.88 %) males. Chronic heart failure (CHF), which is seen in 42 (27.27 %) patients, 101 (65.58 %) did not have CHF and 11 (7.142 %) missing data. Diabetes mellitus (DM) has been observed in 28 (18.18182 %), 118 (76.62 %) without DM, Hypertension has been seen in 118 (76.62 %), 28 (18.18 %) did not have hypertension, chronic kidney disease (CKD) has been seen in 11 (7.14%) patients, 131 (85.064 %) did not have CKD, Chronic obstructive pulmonary disease (COPD) existed in 14 (9.09 %), 132 (85.71 %) did not have COPD. Post myocardial infarction (MI) CHF has been observed in 108 (70.12987%), 42 (27.27 %) did not have CHF. Post MI arrythmia seen in 50 (32.47%) patients, 99 (64.28 %) did not have arrythmia. Early post MI complication such as aneurysm has been seen in 12 (7.79 %), 138 (89.61 %) did not have aneurysm, Dressler syndrome; pericarditis has been seen in 4 (2.59 %) patients, pneumonitis seen in 1 (0.64 %) patient, and pleuritis have not been seen.

**Conclusions:** Systemic manifestations include kidney function impairment or development of new kidney disease. These changes include an increase in the aorta basement diameter, an increase in the right ventricle size, a decrease in the level of serum red blood cells, the serum potassium level, and serum calcium level, with an increase in the serum myoglobin. The high myoglobin level is associated with the development of chronic heart failure. The higher creatinine level and or LDH is associated with arrythmia development. Hypertension is the predominant concomitant disease in victims of MI. And arrythmia is most commonly reported post-MI complication.

**Others:** Females are more frequently affected by post-MI urinary tract infection.

## Introduction

Myocardial infarction (MI) and associated sequalae remain the most reported cause of mortality and morbidity worldwide [1]. The development of cardiovascular events is associated with several modified and non-modified risk factors [2]. Lifestyle changes have a huge impact on the overall health of the cardiovascular system [3, 4]. Unfortunately, sedentary lifestyle is seen in majority of myocardial infarction, which is associated with development of several predisposition factors for cardiovascular events such as dyslipidaemia, insulin resistance, and central obesity or visceral obesity [5–9]. Collectively, these early signs of disturbance of cardiovascular homeostasis are ignored to provoke the progression of dyslipidaemia and insulin resistance to reach enough levels to induce the emergence of cardiovascular events such as MI [10, 11].

Assessment of the early post-MI changes is of huge impact on the further therapy plan and the prognosis of MI victims. One of the early specific markers of myocardial infarction is the elevation of cardiac lactate dehydrogenase 1 and troponins, including troponins I and T. However, systemic changes are rarely assessed in early pot-MI patients [12]. The role of these changes is unclear in determining the prognosis of patients with MI and the potential risk of developing complications such as renal failure. In the current retrospective analysis, we covered the potential role of these systemic and the regularly checked cardiac markers in the prognosis determination and prognosis of the patients [5, 8, 20–23, 9, 13–19].

## Materials and methods

A retrospective cohort analysis for 154 patients for the period 2014-2019. The data has been collected from the republican rehabilitation hospital (history of the patients from the other hospitals are collected in the rehabilitation hospital, that is why we collected the data from her). All the presented data are after first contact with the patient and shows the peak values of post MI changes. Consent from patients has been taken for the collection, analysis, and publication the material and any associated photos or graphs. The missing data have been treated as absent and removed from the analysis. All measurement units are based on the local units used in the laboratories. There was no access to the 2020–2022 year data because of the COVID-19 issues. Pleuritis was not seen in patients and been excluded from the complications report. The presented t-test results are all statistically significant, and other results are not statistically significant at p<0.05. The level of leukocytes and red blood cell level is always mentioned without *10^9^ or *10^12^, respectively. We could not assess the risk hazard for the estimated parameters and the survival rate, limitations of the study.

The T test, the Pearson correlation coefficient and ROC analysis have been used using the Statistica 12 programme (StatSoft, Inc. (2014). STATISTICA (data analysis software system), version 12. www.statsoft.com.).

## Results

The sample included 154 patients with a history of fresh myocardial infarction. Of 154, 51 (33.11%) were female and 103 (66.88%) were male. In the sample, 131 (85.064 %) live in the city, 17 (11.038 %) live in the village, and 5 (3.24 %) live in the town.

Majority of the patient had a concomitant disease prior to their myocardial infarction. 132 (85.71 %) had a concomitant disease, 18 (11.68 %) did not have a concomitant disease, and 4 (2.59 %) missing data. Of the concomitant diseases, chronic heart failure (CHF), which is seen in 42 (27.27 %) patients, 101 (65.58 %) did not have CHF, and 11 (7.14 %) missing data. Diabetes mellitus (DM) has been observed in 28 (18,18 %), 118 (76.62 %) without DM, and 8 (5.19 %) missing data. Hypertension has been observed in 118 (76.62 %), 28 (18.18182%) who did not have hypertension, and 8 (5.19 %) missing data. Chronic kidney disease (CKD) has been observed in 11 (7.14 %) patients, 131 (85.064 %) did not have CKD, and 12 (7.79 %) were missing data. Chronic obstructive lung disease (COPD) existed in 14 (9.09 %), 132 (85.71 %) did not have COPD and 8 (5.19 %) missing data. A disrupted lipid profile was present in 21 (13.63 %), 121 (78.57 %) did not have dyslipidaemia and 12 (7.79 %) missing data. Varicose vein is coexisted in 3 (1.94 %), 137 (88.96 %) did not have varicose vein, and 14 (9.09 %) missing data.

In terms of complications after myocardial infarction, 145 (94.15 %) patients had complication, 5 (3.24 %) did not have complication, and 4 (2.59 %) had missing data. Post-myocardial infarction (MI) CHF has been observed in 108 (70.12 %), 42 (27.27 %) did not have CHF, and 4 (2.59 %) missing data. Post MI arrythmia seen in 50 (32.47 %) patients, 99 (64.28 %) did not have arrythmia, 5 (3.24 %) missing data. Early post MI complication such as aneurysm has been seen in 12 (7.79221%), 138 (89.61 %) did not have aneurysm, and 4 (2.59740%) missing data. Late post MI complications including Dressler syndrome including the pericarditis has been seen in 4 (2.59 %) patients, 146 (94.80 %) did not have pericarditis, and 4 (2.59 %) missing data. Additional components of Dressler syndrome include pneumonitis, which is seen in 1 (0.64 %) patient, 148 (96.10 %) did not have pneumonitis, and 5 (3.24 %) missing data. Whereas, the third component of the Dressler syndrome, pleuritis, have not been seen, therefore, it has not been reported.

Interestingly, 43 (27.92 %) patients had re-infarct, 107 (69.48052 %) were for the first time, and 4 (2.59 %) missing data. Evaluation of the activity level of tissue-type plasminogen activator (t-PA) through serum level, it was elevated in 7 (4.54 %) patients, 5 (3.24 %) in the normal range, and other 142 (92.20 %) did not measure their d-dimer level.

The primary descriptive statistics demonstrated in (*Table*) 1.

**Table 1:** The primary descriptive statistics of the sample. Abbreviations: EF; ejection fraction, EDV; end diastolic volume, LV; left ventricle, LA; left atrium, RV; right ventricle, RA; right atrium, Hb; hemoglobin, RBC; red blood cell, ESR; erythrocyte sedimentation rate of erythrocytes, CKP-MB; creatinine phosphokinase-MB fraction, LDH; lactate dehydrogenase, ALT; alanine transaminase, AST; aspartate transaminase, LDL; low density lipoprotein, TAG; triacyl glycerides, APTT; activated prothrombin time, PT; prothrombin time.

| Variable | Descriptive Statistics |  |  |  |  |  |
| --- | --- | --- | --- | --- | --- | --- |
|  | Valid N | % Valid obs. | Mean | Median | Minimum | Maximum |
| Hospitalization days | 153 | 99.35 | 14.53 | 15.0 | 0.00 | 24 |
| Age (years) | 152 | 98.70 | 61.64 | 62.5 | 30.00 | 86 |
| Body weight (Kg) | 31 | 20.12 | 82.26 | 85.000 | 62.00 | 112 |
| Height (cm) | 31 | 20.12 | 170.58 | 172.00 | 155.0 | 185 |
| BMI (Kg/m<sup>2</sup>) | 31 | 20.12 | 28.56 | 27.88 | 21.79 | 41 |
| HR (beat/mint) | 148 | 96.10 | 79.82 | 78.00 | 53.00 | 130 |
| SBP (mmHg) | 148 | 96.10 | 139.57 | 140.00 | 60.00 | 260 |
| DBP (mmHg) | 148 | 96.10 | 84.05 | 80.00 | 40.00 | 130 |
| Aorta diameter at the Valsalva sinuses (mm) | 144 | 93.50 | 35.21 | 35.00 | 20.00 | 46 |
| EF (%) | 142 | 92.20 | 54.55 | 55.000 | 25.00 | 80 |
| EDV (ml) | 113 | 73.37 | 134.30 | 129.00 | 62.00 | 234 |
| End diastolic LV size (mm) | 132 | 85.71 | 52.35 | 51.150 | 10.00 | 89 |
| End diastolic LA size (mm) | 133 | 86.36 | 40.09 | 40.00 | 28.00 | 62 |
| End diastolic RV size (mm) | 144 | 93.50 | 26.75 | 26.25 | 5.00 | 44 |
|  | Valid N | % Valid obs. | Mean | Median | Minimum | Maximum |
| End diastolic RA size (mm) | 130 | 84.41 | 35.73 | 36.00 | 24.00 | 49 |
| Hb | 143 | 92.85 | 137.10 | 138.0 | 85.00 | 170 |
| Leukocytes (/L) | 141 | 91.55 | 8.82 | 8.30 | 2.80 | 18 |
| RBC (/L) | 118 | 76.62 | 5.12 | 4.50 | 2.60 | 24 |
| Lymphocytes (%) | 130 | 84.41 | 25.51 | 25.50 | 2.00 | 54 |
| Thrombocytes (/L) | 111 | 72.07 | 206.76 | 200.00 | 83.00 | 506 |
| ESR (mm/hr) | 138 | 89.61 | 19.04 | 15.50 | 2.00 | 60 |
| Urine ketones (mmol/l) | 1 | 0.64 | 0.50 | 0.50 | 0.50 | 1 |
| Urine proteins (mmol/l) | 137 | 88.96 | 5.27 | 0.00 | 0.00 | 150 |
| Urine leukocytes (microscopically) | 142 | 92.20 | 2.64 | 1.00 | 0.00 | 50 |
| Urine epithelial cells (microscopically) | 136 | 88.31 | 1.86 | 1.000 | 0.00 | 20 |
| Troponin I (ng/ml) | 126 | 81.81 | 4.74 | 1.20 | 0.00 | 33 |
| Myoglobin (ng/ml) | 3 | 1.94 | 111.30 | 71.0 | 63.00 | 200 |
| CKP-MB 1 (U/l) | 129 | 83.76 | 97.01 | 29.0 | 6.00 | 2881 |
| CKP-MB 2 (U/l) | 71 | 46.10 | 79.26 | 42.0 | 6.00 | 508 |
| LDH 1 (U/l) | 108 | 70.12 | 652.30 | 475.5 | 30.00 | 3900 |
| LDH 2 (U/l) | 56 | 36.36 | 21640.80 | 492.0 | 97.00 | 1177 |
| INR (Sec) | 125 | 81.16 | 1.25 | 1.00 | 0.80 | 18 |
| Total bilirubin (mmol/l) | 108 | 70.12 | 16.53 | 17.68 | 4.42 | 36 |
| ALT (mmol/l) | 140 | 90.90 | 42.65 | 28.00 | 1.19 | 361 |
| AST (mmol/l) | 140 | 90.90 | 55.57 | 30.00 | 5.10 | 423 |
| Total cholesterol (mmol/l) | 141 | 91.55 | 5.23 | 5.10 | 2.40 | 9 |
| LDL (mmol/l) | 138 | 89.61 | 23.20 | 3.24 | 0.92 | 2766 |
|  | Valid N | % Valid obs. | Mean | Median | Minimum | Maximum |
| TAG (mmol/l) | 140 | 90.90 | 3.65 | 1.56 | 0.27 | 138 |
| Na (mmol/l) | 122 | 79.22 | 139.76 | 142.75 | 1.76 | 151 |
| K (mmol/l) | 126 | 81.81 | 6.02 | 4.06 | 1.00 | 92 |
| Creatinine (μmol/L) | 142 | 92.20 | 81.56 | 84.00 | 0.00 | 176 |
| Ca (mmol/l) | 115 | 74.67 | 1.69 | 1.18 | 0.25 | 38 |
| Urea (mmol/l) | 139 | 90.25 | 7.79 | 6.70 | 2.50 | 93 |
| Glucose (mmol/l) | 135 | 87.66 | 6.86 | 6.10 | 4.00 | 19 |
| APTT (Sec) | 74 | 48.05 | 27.31 | 25.50 | 15.00 | 76 |
| Fibrinogen (ng/ml) | 117 | 75.97 | 3386.85 | 3500.00 | 300.00 | 7500 |
| PT (Sec) | 1 | 0.64 | 94.00 | 94.00 | 94.00 | 94 |

Males are affected by myocardial infarction earlier than females; the cut-off point for age is 67 years old. (*Figure 1*)

**Figure 1:**
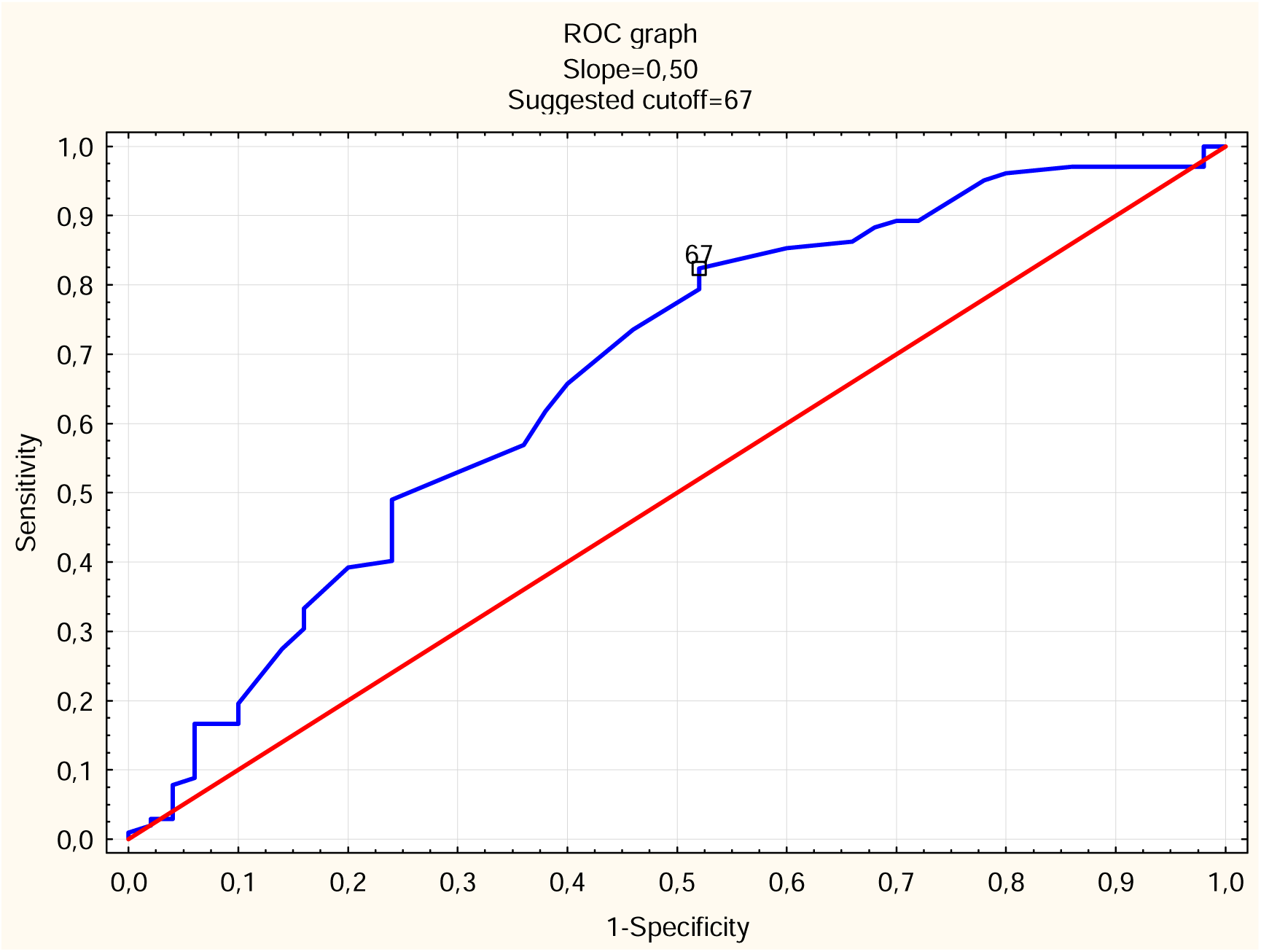
Cut-off point for the age of the patients divided into male and female. Males are less than 67 years old. However, the female affected at more than 67 years of age.

In male group the mean age is 59.76 years old and in female group the mean age is 65.46 years old, which is statistically significant difference, t value 3.30, p<0.001. Furthermore, the mean height of the male is 172.57 cm and in female 164.88 cm, which is a statistically significant difference, t value -3.26, p< 0.002. On echocardiogram, in male group, the mean diameter of the aorta is 36.23 mm, in female group, the mean diameter of the aorta is 33.04 mm, which is statistically significant difference, t value -4.24, p<0.00003. In male group, the mean right atrium size on echocardiogram equal to 36.26 mm, in female group equal to 34.56 mm, which is statistically significant difference, t value -2.06, p< 0.04. The mean hemoglobin level in men was 140.26 and in women 130.66, which is a statistically significant difference, t value -3.71, p<0.0002. In urine analysis, in male group, mean number of leukocytes in urine was 1.52 under microscope and in female group was 4.91, which is statistically significant difference, t value 3.11, p< 0.002. Furthermore, in urine analysis, the number of epithelial cells in the male 1.36 under microscope and in the female group 2.91, which is a statistically significant difference, t value 3.12, p< 0.002. In the male group, the mean myoglobin level was 67.00 ng/ml and in the female group 199.90 ng/ml, which is a statistically significant difference, t value 19.18, p< 0.03. (*Table* 2)

**Table 2:**
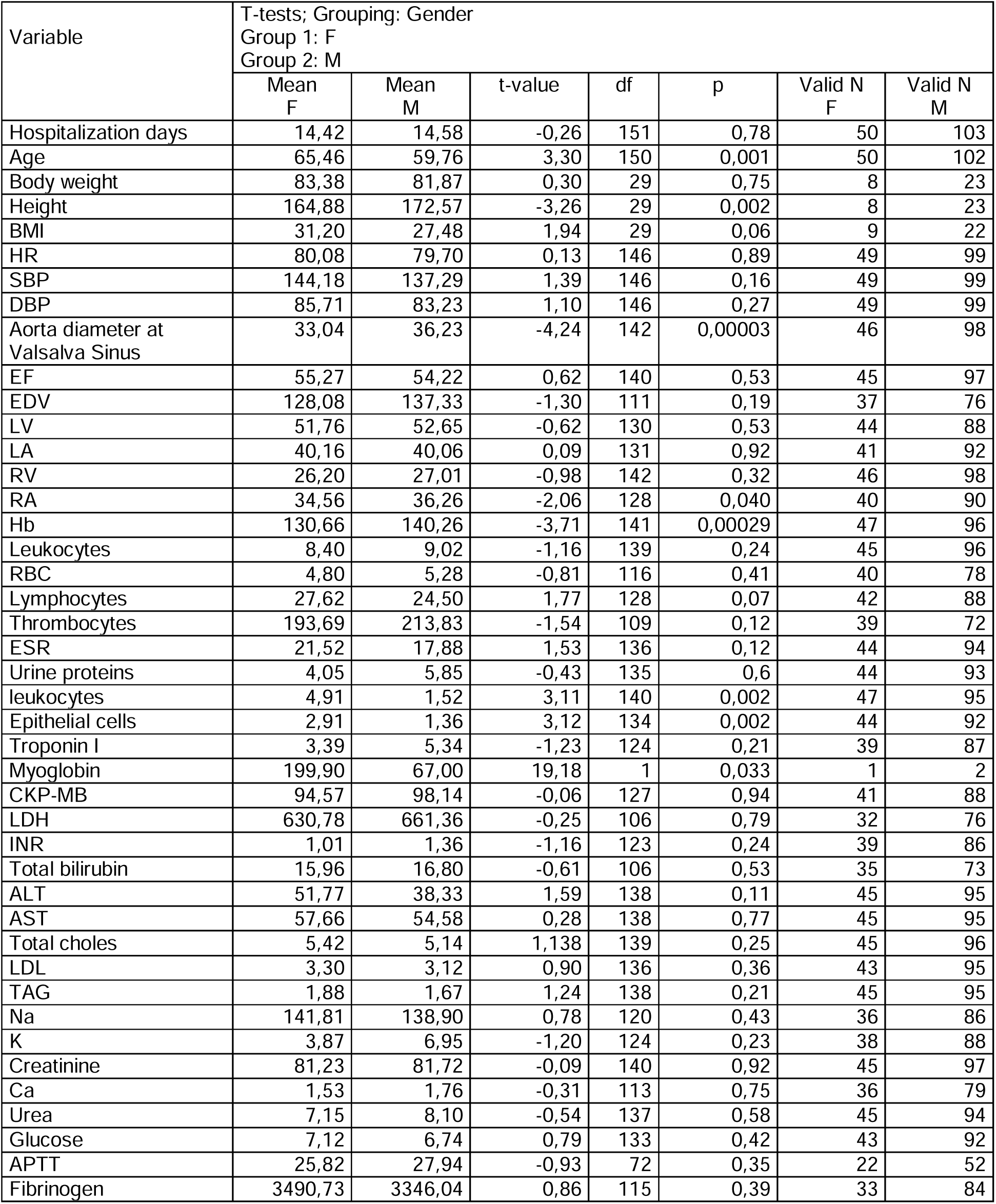
Gender role in the numerical variables.

Patients with concomitant diseases have a mean heart rate (HR) equal to 80.82 beats / minute and patients without concomitant disease have a mean HR equal to 72.39 beats / minute, which is a statistically significant difference, t value 2.05, p< 0.04. Furthermore, patients with concomitant diseases have a mean diastolic size of the right ventricle is 27.13 mm and patients without concomitant disease have a mean diastolic size of the right ventricle is 23.85 mm, which is a statistically significant difference, t value 2.79, p< 0.005. Moreover, patients with concomitant disease have a mean protein in urine 3.18 mg/l, and the patients without concomitant disease had a mean level of urine protein 21.25 mg/l, which is statistically significant difference, t value -3.10, p< 0.002. Interestingly, patients with concomitant disease had lower calcium level a mean equal to 1.31 mmol/l and the patients without concomitant disease had a mean calcium level 3.84 mmol/l, which is statistically significant difference, t value -2.67, p< 0.008. (*Figure 2*)

**Figure 2:**
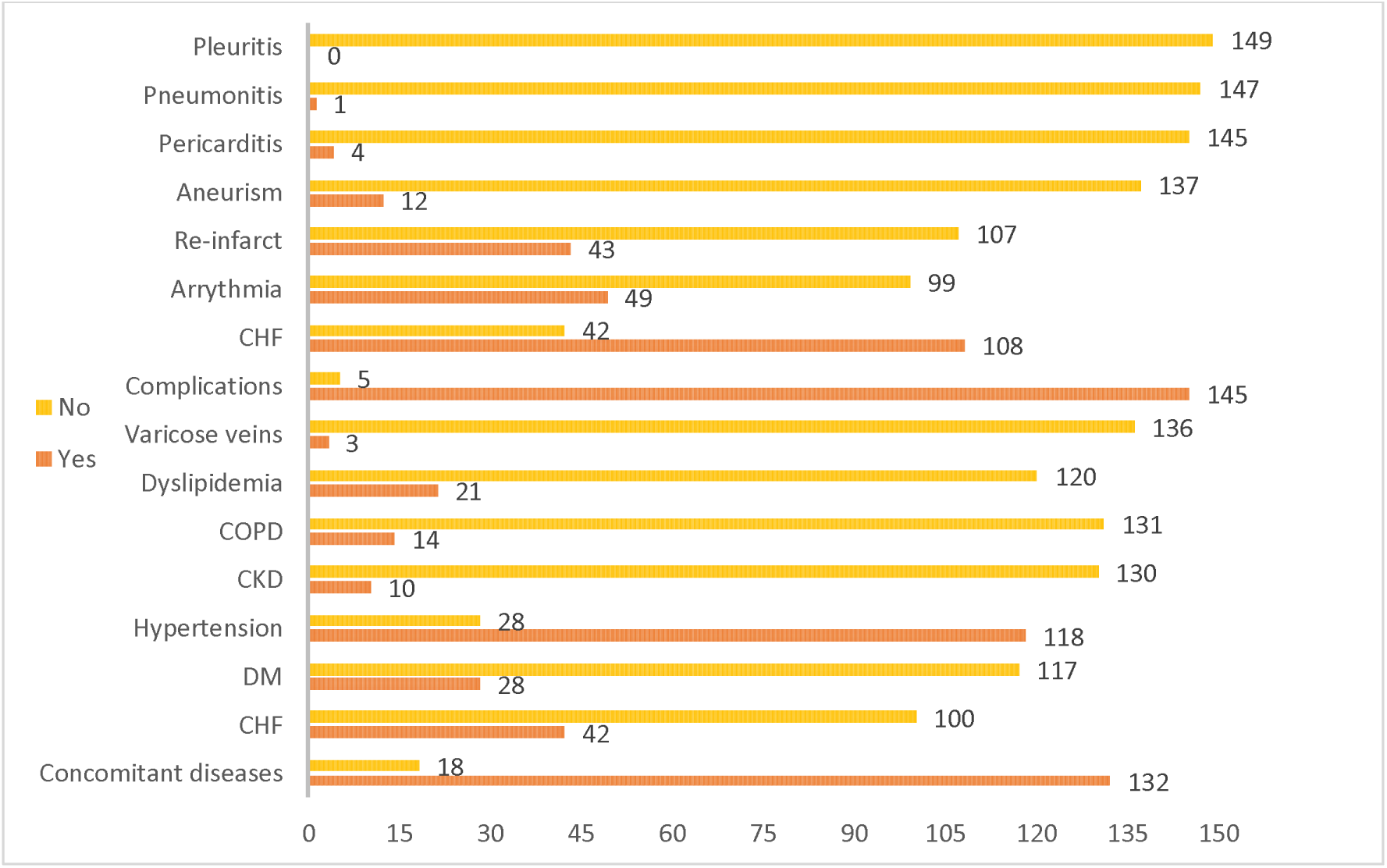
Diagramed presentation of the concomitant and the post MI compilations. From below complications (CHF, DM, hypertension, CKD, COPD, dyslipidaemia, varicose veins) belong to concomitant diseases before the development of MI. Abbreviations: CHF; chronic heart failure, COPD; chronic obstructive pulmonary disease, CKD; chronic kidney disease, DM; diabetes mellites.

Patients with CHF have a mean thrombocyte level equal to 185.26 / l, while patients without CHF have a mean thrombocyte level of 216.24 /l, which is a statistically significant difference, t value - 2.15, p< 0.03.

Mean glucose level in diabetic patients equal to 9.93 mmol/l and in non-diabetic patients equal to 6.13 mmol/l, which is statistically significant difference, t value -7.97, p< 0.0001. The coexistence of DM in patients increases the mean diastolic size of the left ventricle 55.60 mm, while, in patients without DM, the mean diastolic size of the left ventricle to equal 51.42 mm, which is a statistically significant difference, t value -2.53, p< 0.012. The mean sedimentation rate (ESR) in patients with DM equals 23.60 mm/hr, and equal to 17.43 mm/hr in non-diabetic patients, which is statistically significant difference, t value -2.22, p< 0.027. The mean creatinine phosphokinase-MB fraction is dramatically higher in patients with DM 233.36 U/L and lower in non-diabetic patients 66.05 U / L, which is a statistically significant difference, t value --2.62, p< 0.009. Diabetic patients experience a high level of triacyl glyceride (TAG) 2.17 mmol / l, while non-diabetic patients had a lower TAG level of 1.66 mmol/l, which is a statistically significant difference, t value -2.56, p< 0.011.

Hypertensive patients have a low mean level of red blood cells (RBC) level 4.55 /l in compare to non-hypertensive patients 7.7, which is statistically significant difference, t value -4.63276, p< 0.000010. In hypertensive patients, the mean LDH 1 is 608.21 U/l, INR is 1.05, sodium is 141.70 mmol/l, potassium is 4.04 mmol/l, and calcium is 1.14 mmol/l, whereas, in non-hypertensive patients, the mean level of LDH 1 is 908.8 U/l, INR is 2.1, sodium is 130.6 mmol/l, potassium is 14.8 mmol/l, calcium is 3.9 mmol/l, which is statistically significant difference; t value -2.07, -3.04, 2.51, -3.62, -3.19; p< 0.04, 0.0028, 0.013, 0.0004, 0.001, respectively.

CKD patients have a high mean troponin I level equal to 22.70 ng / ml compared to non-CKD patients 3.72 ng/ml, which is a statistically significant difference, t value -6.84, p< 0.0001. Also, CKD patients have higher mean level of CPK-MB fraction equal to 475.61 U/l in compare to non-CKD patients equal to 78.38 U/l, which is statistically significant difference, t value -3.63, p< 0.0004. The mean creatinine level is higher in patients with CKD at 109.81 µmol/L comparing to non-CKD patients, which is a statistically significant difference, t value -3.63, p< 0.0004.

Interestingly, patients with COPD have a mean diastolic size of the left ventricle is 48.46 mm, which is less than those of patients 52.73 mm, which is a statistically significant difference, t value 1.99, p< 0.048. However, patients with COPD have a mean diastolic size of the right atrium is 39.55 mm compared to patients 35.41 mm, which is a statistically significant difference, t value -3.06, p< 0.002. Furthermore, COPD patients have a mean level of RBC equal to 145.29 compared to non-COPD patients 136.39, which is a statistically significant difference, t value -2.11, p< 0.03.

Patients with dyslipidemia have mean troponin I 9.4 ng/ml, whereas, patients without dyslipidemia have mean troponin I 3.64 ng/ml, which is statistically significant difference, t value -2.99, p< 0.003. Additionally, patients with dyslipidaemia have mean total bilirubin 19.3 mmol / l and 15.72 mmol/l in patients without dyslipidaemia, which is a statistically significant difference, t value -2.12, p< 0.03. Furthermore, patients with dyslipidaemia have a mean serum sodium level of 132.3 mmol/l and a mean serum potassium level of 8.4 mmol / l, while in patients without dyslipidaemia, the mean sodium serum level is 142.24 mmol/l and a mean serum potassium level of 4.07 mmol/l, which is a statistically significant difference; t value 2.74, -2.09; p< 0.006, 0.03.

The mean leukocytes level in patients with varicose veins is 12.07 /l, whereas, in patients without varicose veins, the mean leukocytes level is 8.71 /l, which is statistically significant difference; t value -1.99, p< 0.04. The mean RBC level in patients with varicose veins is 9.87 /l, whereas, in patients without varicose veins, the mean RBC level is 4.56 /l, which is statistically significant difference; t value -6.88, p< 0.00. The mean level of ESR in patients with varicose veins is 37.67 mm / h, while, in patients without varicose veins, the mean ESR level is 18.25 mm / h, which is statistically significant difference; t value -2.64, p< 0.009.

In terms of complications, patients with complications have been hospitalized for 14.65 days, whereas, patients without complications hospitalized for 11.40 days, which is statistically significant difference; t value 2.05, p< 0.04. In patients with complications, the mean aorta diameter 35.38 mm, whereas, in patients without complications, the mean aorta diameter 29.25 mm, which is statistically significant difference; t value 2.77, p< 0.006. In patients with complications, the mean diastolic size is of the right ventricle is 26.89 mm, while, in patients without complications, the mean diastolic size of the right ventricle 21.25mm, which is a statistically significant difference; t 2.44, p< 0.01. In patients with complications, the mean level of RBC 4.87 / l, while in patients without complications, the mean level of RBC 12.40 /l, which is a statistically significant difference; t value -5.37, p< 0.0001. In patients with complications, in the general urine analysis, microscopically, the mean level of urea proteins 4.30, while, in patients without complications, the mean level of urea proteins 50.00, which is a statistically significant difference; t -3.61, p< 0.0004. In patients with complications, the mean potassium level 5.37 mmol/l and mean calcium level 1.58 mmol/l, whereas, in patients without complications, the mean potassium level 32.99 mmol/l and mean calcium level is 5.99 mmol/l, which is statistically significant difference; t value -3.75, -2.05; p< 0.0002, 0.04, respectively.

Patients with post-MI CHF have mean myoglobin level 199.90 ng/ml, whereas, in patients without post-MI CHF, the mean myoglobin level 67.00 ng/ml, which is statistically significant difference; t value 19.18, p< 0.033.

Patients with a history of previous infarction have creatinine 90.48 µmol/l in compare to patients without reinfarction 78.01 µmol/l, which is a statistically significant difference; t value 2.26, p< 0.02

The mean age of patients with arrythmia is 65.58 years, while the mean age of patients without arrythmia is 59.68 years, which is statistically significant difference; t value 3.32, p< 0.001. The mean height of patients with arrythmia is 173.67 cm, while the mean height of patients without arrythmia is 168.61 cm, which is statistically significant difference; t value 2.23, p< 0.03. The mean ejection fraction (EF) of patients with arrythmia is 50.40 %, while the mean EF of patients without arrythmia is 56.43 %, which is statistically significant difference; t -3.81, p< 0.0002. The mean diastolic size of the left atrium of patients with arrythmia is 42.14 mm, while the mean diastolic size of the left atrium of patients without arrythmia is a 39.12 mm, which is statistically significant difference; t 3.00, p< 0.003. The mean diastolic size right atrium of patients with arrythmia is 37.36 mm, whereas, the mean diastolic size of the right atrium of patients without arrythmia is 34.95 mm, which is statistically significant difference; t value 2.96, p< 0.003. The mean CKP-MB of patients with arrythmia is 181.82 U / l, while the mean CKP-MB of patients without arrythmia is 56.68 U/l, which is statistically significant difference; t 2.34, p< 0.02. The mean serum lactate dehydrogenase (LDH) level of patients with arrythmia is 888.37 U/l, whereas, the mean lactate dehydrogenase level of patients without arrythmia is a 533.49 U/l, which is statistically significant difference; t value 3.16, p< 0.002. The mean serum level of aspartate transaminase of patients with arrythmia is 71.53 mmol / l, while the mean lactate dehydrogenase serum level of patients without arrythmia is a 47.38 mmol/l, which is statistically significant difference; t value 2.23, p< 0.026.

The mean EF of patients with aneurysm is 43.00 %, whereas, the mean EF of patients without aneurysm is 55.59 %, which is statistically significant difference; t value 4.57, p< 0.00001. The mean level in patients with aneurysm is 9.70 / l, while the mean leukocyte level of patients without aneurysm is 2.11 /l, which is statistically significant difference; t -3.83, p< 0.0001. The mean epithelial cell level in general urine analysis in patients with aneurysm is 5.40, whereas, the mean epithelial cell level in patients without aneurysm is 1.58, which is statistically significant difference; t value -4.41, p< 0.00002.

The mean diastolic size of the left atrium of patients with pericarditis is 45.5 mm, while the mean diastolic size of the left atrium of patients without pericarditis is 39.90 mm, which is statistically significant difference; t value -2.01, p< 0.04. The mean creatinine level of patients with pericarditis is 52.5 µmol/L, whereas, the mean creatinine level of patients without pericarditis is 82.39 µmol/L, which is statistically significant difference; t value 1.98, p< 0.048.

Patients with pneumonitis have mean aorta diameter of 20.00 mm, whereas, the mean aorta diameter of patients without pneumonitis is 35.32 mm, which is statistically significant difference; t value 3.55, p< 0.0005. Patients with pneumonitis have mean serum leukocytes level of 14.80 /l, whereas, the mean serum leukocytes level of patients without pneumonitis is 8.70 /l, which is statistically significant difference; t value -2.11, p< 0.036. Patients with pneumonitis have a mean ESR level of 48.00 mm / hour, while the mean ESR level of patients without pneumonitis is an 18.47 mm/he, which is statistically significant difference; t value -2.36, p< 0.019. Furthermore, patients with pneumonitis have a mean leukocyte level of 22.00, whereas, the mean urine leukocyte level in urine of patients without pneumonitis is a 2.53, which is statistically significant difference; t value -3.15, p< 0.001. The mean CKP-MB in patients with pneumonitis is 1164.00 U / l, while the mean CKP-MB in patients without pneumonitis is 89.60 U/l, which is statistically significant difference; t -3.92, p< 0.0001.

The mean diameter of the aorta, EF, EDV, troponin I, CKP-MB, and APTT in patients with elevated troponin T is 28.00 mm, 36.00 %, 62.00 ml, 33.00 ng/ml, 2881.00 U / l, and 49.10 s, respectively. Whereas, the mean aorta diameter, EF, EDV, troponin I, CKP-MB, APTT in patients without elevation in troponin T is 35.31 mm, 54.68%, 134.95 ml, 4.51 ng/ml, 75.26 U/l, 27.01 sec, respectively, which is statistically significant difference; t value 2.34, 2.01, 2.07, -3.61, -19.34, -2.55, p< 0.020, 0.045, 0.04, 0.0004, 0.00 0.012, respectively.

The mean age of patients with an elevated D-dimer level is 68.14 years, and the mean age of patients with normal D-dimer level is 56.60 years, which is statistically significant difference; t value 2.25, p< 0.04. The mean diastolic size of the right ventricle of patients with elevated D-dimer level is 25.93 mm, and mean diastolic size of the right ventricle of patients with normal D-dimer is 29.40 mm, which is statistically significant difference; t value -2.35, p< 0.04.

Comparing the dependent variable shows no statistical differences. (*Figure 3*)

**Figure 3:**
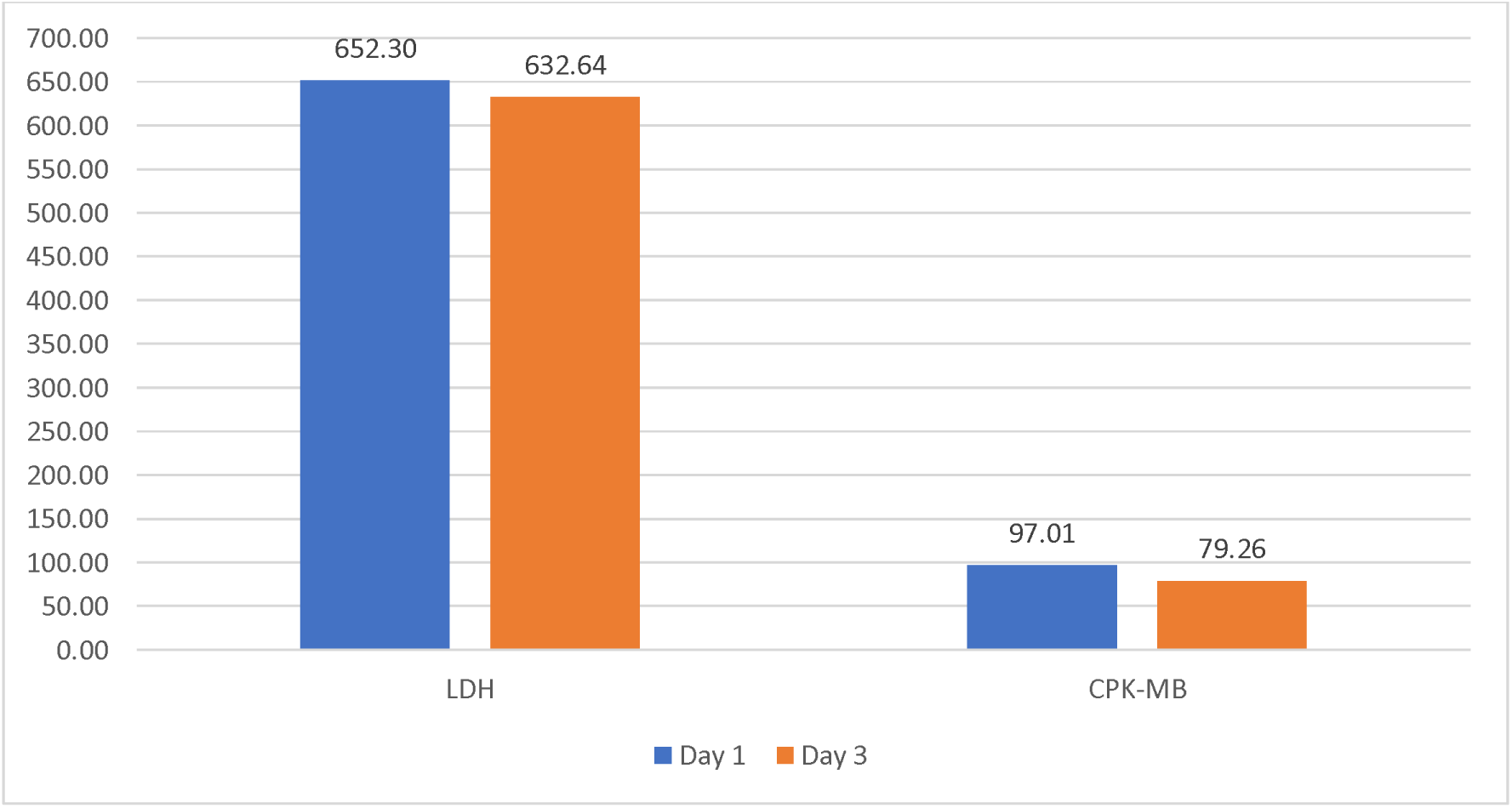
Graphical presentation of the dynamical changes in the levels of LDH (lactate dehydrogenase) and CPK-MB (creatinine phosphokinase-MB fraction). Both LDH and CPK-MB fraction measured by U/L. this analysis conducted between two days for all patients as a follow up after MI.

In terms of correlations, a full table shows the correlation between all the continuous variables is presented. (*Table 3*)

An interesting correlation between the level of LDL and the diameter of the aorta basement has been observed. (*Figure 4.1, 4.2*) It has been shown that when the diastolic size of the left ventricle increases, the EF decreases dramatically. (*Figure 4*)

**Figure 4.1 and 4.2:**
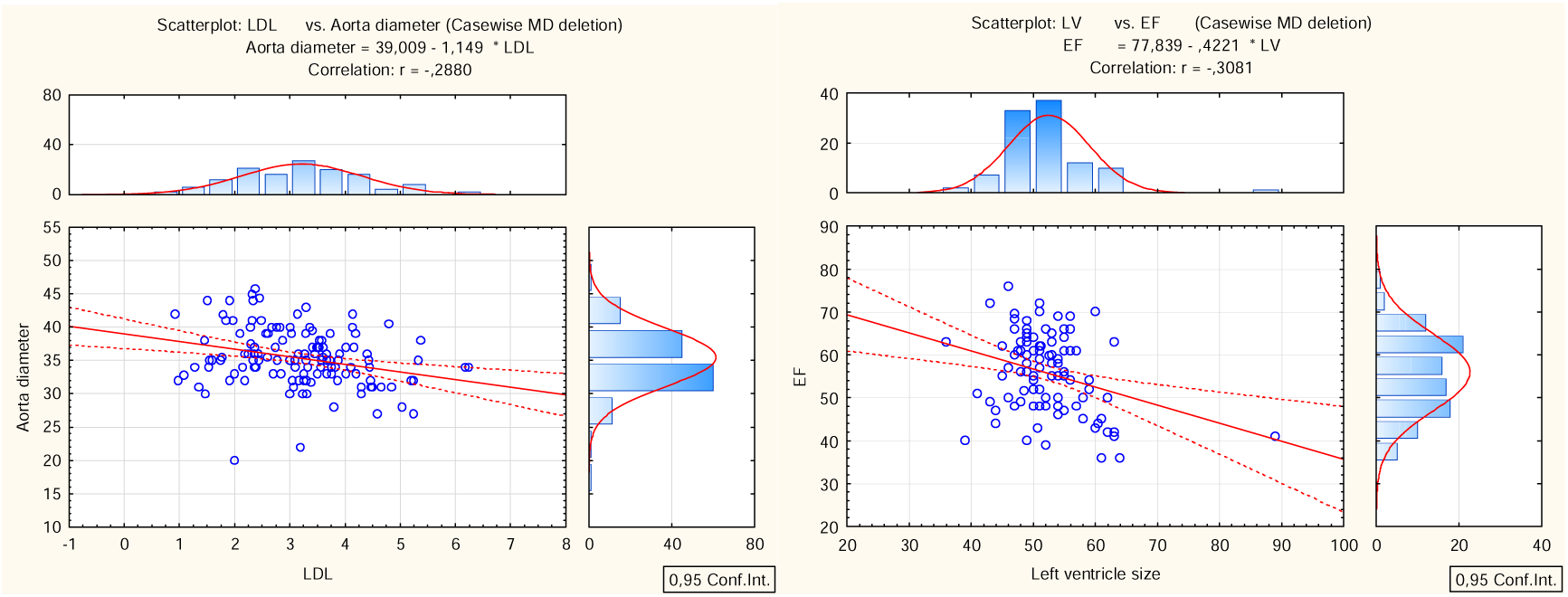
A direct correlation between LDL level and the aorta diameter. Furthermore, an opposite correlation between the left ventricle (LV) diastolic size and the EF.

## Discussion

Post-MI changes include early and late pathophysiological changes, early post-MI complications include ventricular rupture, aneurism, arrythmia, sudden cardiac arrest, and death [24–26]. However, late changes include pericarditis, pneumonitis, pleuritis, and post-infarction cardiosclerosis and dyskinesia [8, 27]. Such manifestations are associated with some laboratory and instrumental changes. Assessment of these changes helps prevent or slow down or at least make a prognosis for patients with MI. The low level of thrombocytes in patients with concomitant CHF returns to the use of antiaggregant and anticoagulants for a long time.

The high level of leukocytes in diabetic patients endorses the hypothesis of a mild chronic mild inflammatory syndrome in patients with metabolic syndrome [9, 21, 23, 28]. The high TAG values in diabetic patients are due to lipogenesis from high glucose serum levels and metabolic disorders associated with chronic lipid peroxidation and biogenesis disorders [28, 29].

The dramatic increase in diastolic size in of the right ventricle patients with COPD is an indicator of the stable stage of pulmonary heart, which is associated with pulmonary hypertension and blood stagnation in the pulmonary circulation [30, 31]. Typically, these patients experience coughing and shortness of breath in the background of distress syndrome, fluid accumulation in pulmonary tissue, and impaired ability of pneumocytes to exchange gas. As compensation for respiratory hypoxemia, the elevation of the levels of erythropoietin and the further elevation of erythropoiesis, which is demonstrated in the high level of Hb in patients with COPD.

Leukocytosis is seen in patients with varicose veins and is an indicator of persistent inflammation with an increase in tissue hypoxia and a further increase in the hypoxia inducer factor to stimulate erythropoietin production and an additional increase in the RBC level of red blood cells in patients with varicose veins [32, 33]. This persistent inflammation is confirmed by the elevation of the ESR level in patients with varicose veins [34–37].

Post-MI complications reported in 4/5 of the sample and these complications associated with some laboratory and instrumental changes. Suggesting that post MI laboratory changes are not limited to the local myocardial infarction changes to involves systemic manifestations such as kidney function impairment or development of new kidney disease. These changes include an increase in the aorta diameter at the basement level, an increase in the right ventricle wall, a decrease in the level of RBC, the serum potassium level, and serum calcium level, with an increase in the serum myoglobin. High myoglobin level is associated with the development of chronic heart failure.

Re-infarct worsens the kidney function and induces renal failure [38–41]. Aging is a risk factor for MI and arrythmia development. Arrythmia increases CPK-MB, LDH, AST, diastolic size of both atriums, and decreases EF. The higher the troponin level, the lower the EF and the fewer further sequalae.

An early post-MI aneurysm significantly alters cardiac output and further reduces systemic systolic blood pressure. These changes are associated with the reduction in the blood flow in to the renal afferent fibers, which are sensed by the juxtaglomerular apparatus that leads to activation the renin angiotensin aldosterone system with the anti-diuretic hormone. Therefore, a reduction in kidney function is observed in patients with MI. Additionally, elevation of creatinine in aneurysm patients is typical for impaired kidney function particularly filtration due to hypoperfusion of the glomeruli. The higher creatinine level is associated with arrythmia development. Also, the higher LDH is associated with arrythmia development.

Patients with pneumonitis are having a high leukocyte in serum and in the urine, high ESR, and high CKP-MB fraction. However, these patients experience small diameter of the aorta basement.

## Conclusions

The early prognostic value of the instrumental and laboratory changes is complication dependent. With age, the level of D-dimer and thrombocytes increases. The higher the troponin level, the lower the EF and the fewer further sequalae. Systemic manifestations include kidney function impairment or development of new kidney disease. These changes include an increase in the aorta diameter at the basement level, an increase in the right ventricle wall, a decrease in the level of serum red blood cells, the serum potassium level, and serum calcium level, with an increase in the serum myoglobin. The higher creatinine level is associated with arrythmia development. Also, the higher LDH is associated with arrythmia development.

## Data Availability

All data produced in the present study are available upon reasonable request to the authors

## List of abbreviations

CHF: chronic heart failure,
DM: Diabetes mellitus,
CKD: Chronic kidney disease,
COPD: Chronic obstructive pulmonary disease,
MI: myocardial infarction,
HR: heart rate,
ESR: erythrocytes sedimentation rate,
RBC: red blood cells

## Declarations

### ETHICS APPROVAL AND CONSENT TO PARTICIPATE

The study approved by the National Research Mordovia State University6 Russia, from “Ethics Committee Requirement N8/2 from 30.06.2022”.

### HUMAN AND ANIMAL RIGHTS

No animals were used in this research. All human research procedures followed were in accordance with the ethical standards of the committee responsible for human experimentation (institutional and national), and with the Helsinki Declaration of 1975, as revised in 2013.

### CONSENT FOR PUBLICATION

Written informed consent was obtained from the participants for publication of study results and any accompanying images.

### AVAILABILITY OF DATA AND MATERIALS

Not applicable.

## ACKNOWLEDGEMENTS

Declared none.

## AUTHORS’ CONTRIBUTIONS

MB collected the data from the hospital, inserted them into the excel file, analyzed the statistical data, wrote the draft, and revised the final version of the paper. NB, VG, and EV inserted data into the excel file. EG gave me access to the rehabilitation hospital to collect the data. All authors have read and approved the manuscript.

---

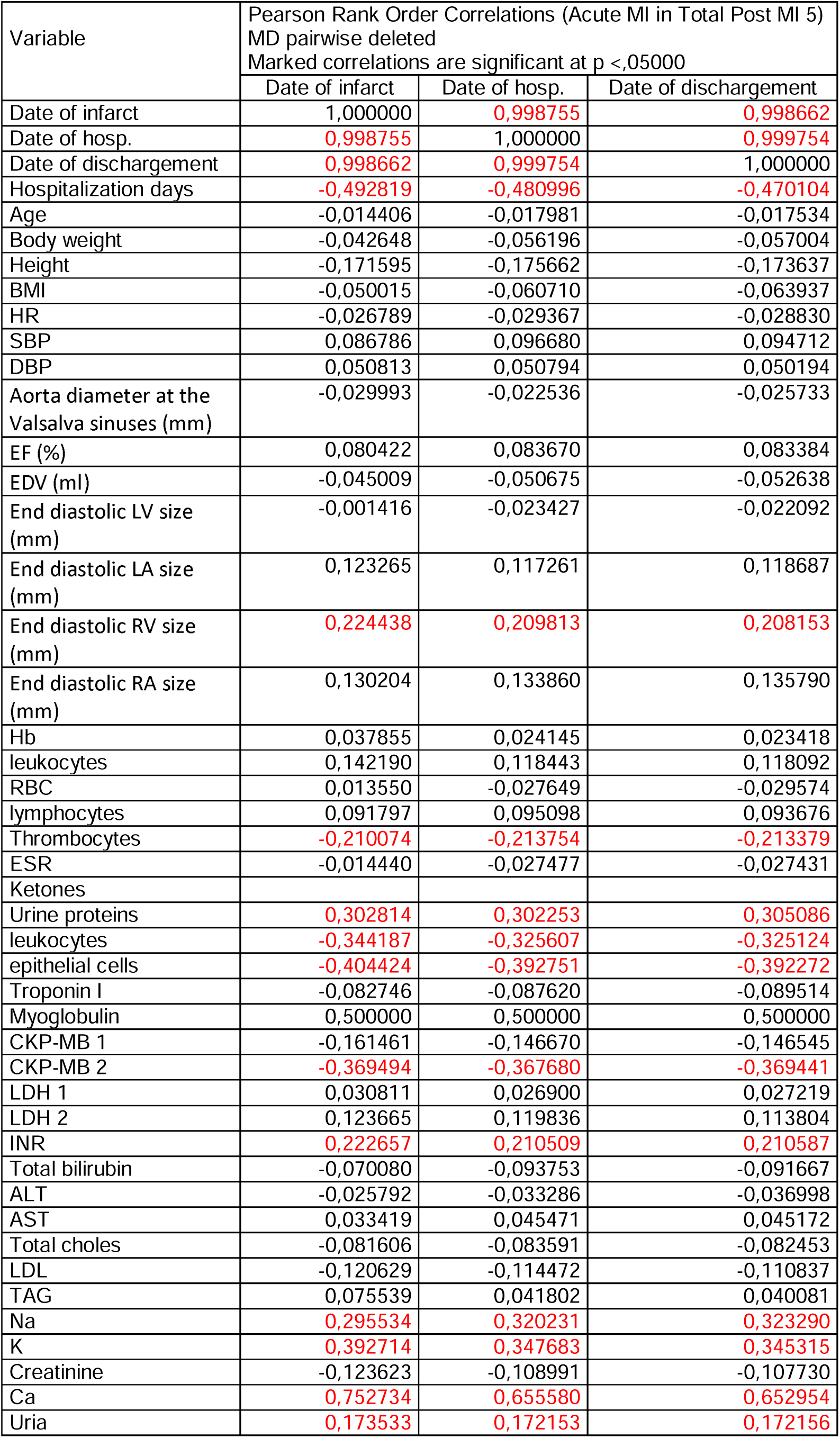
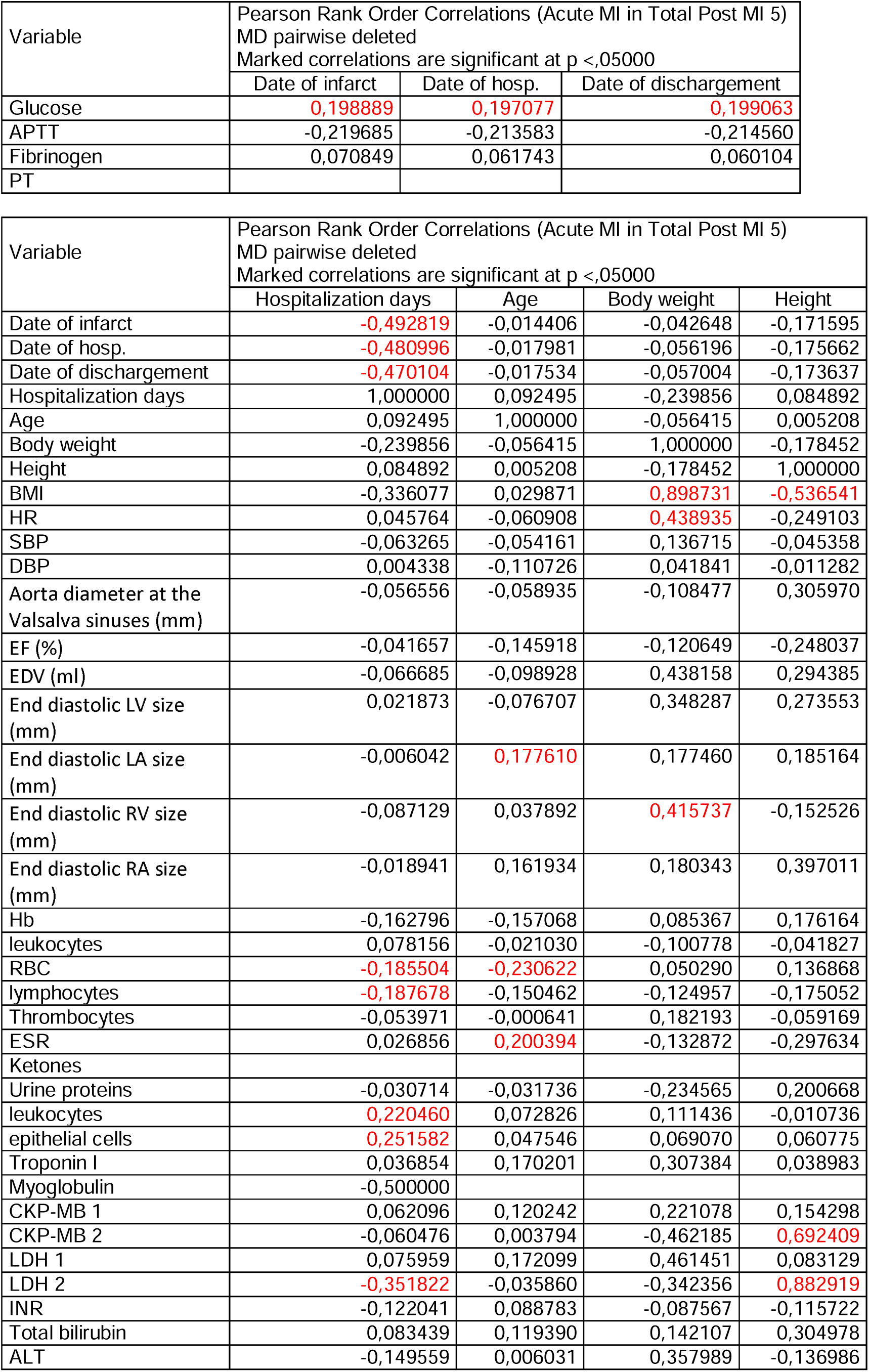
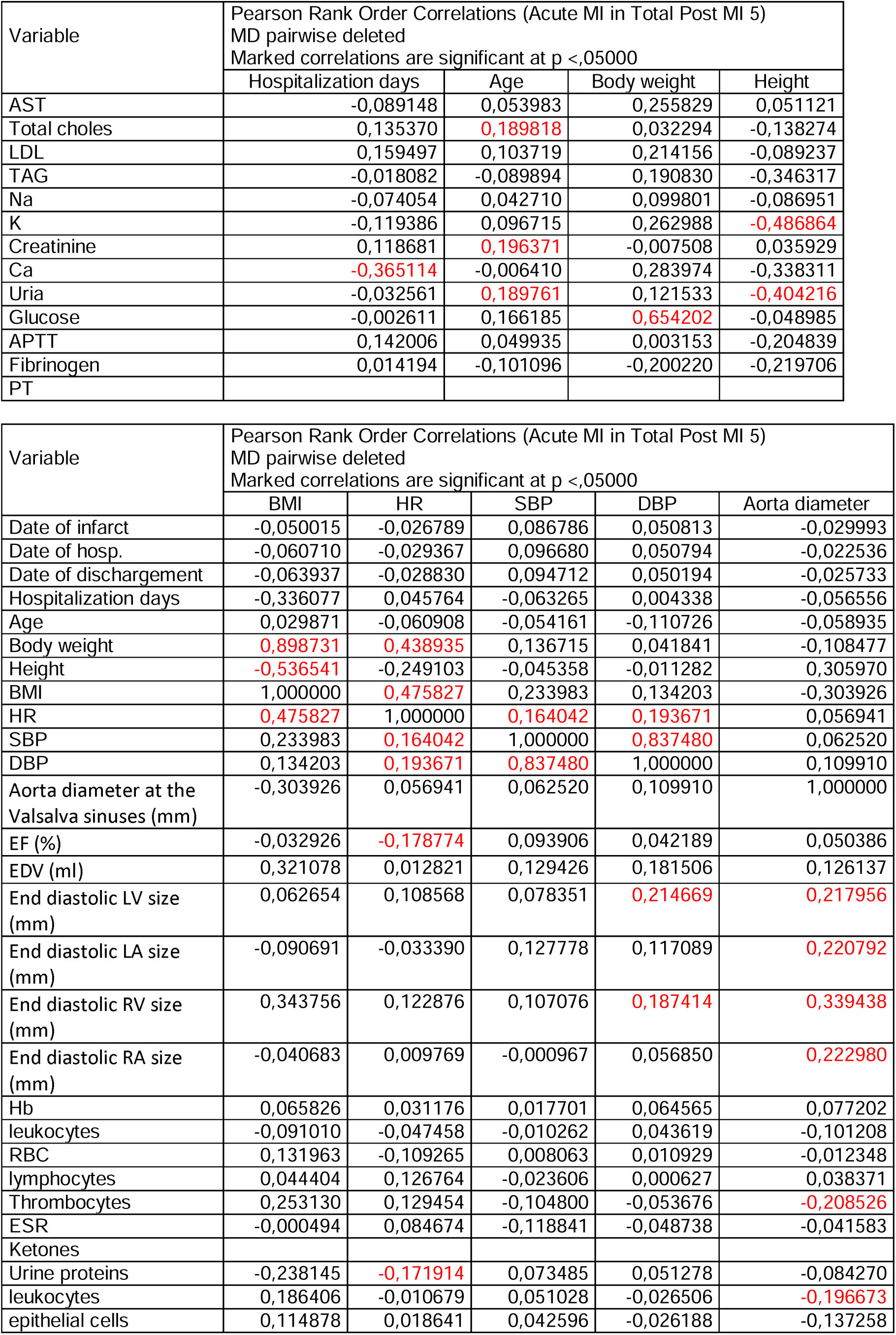
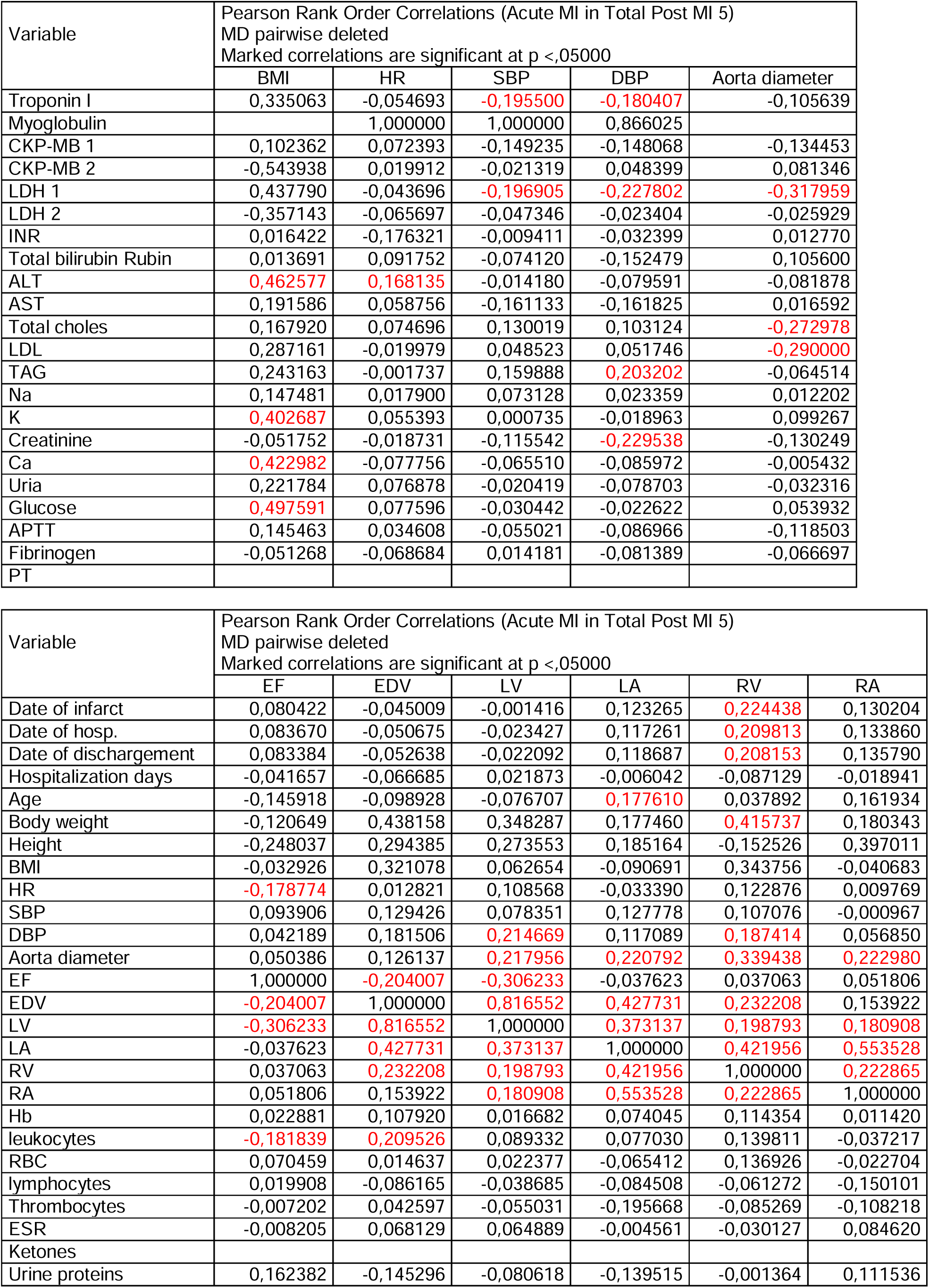
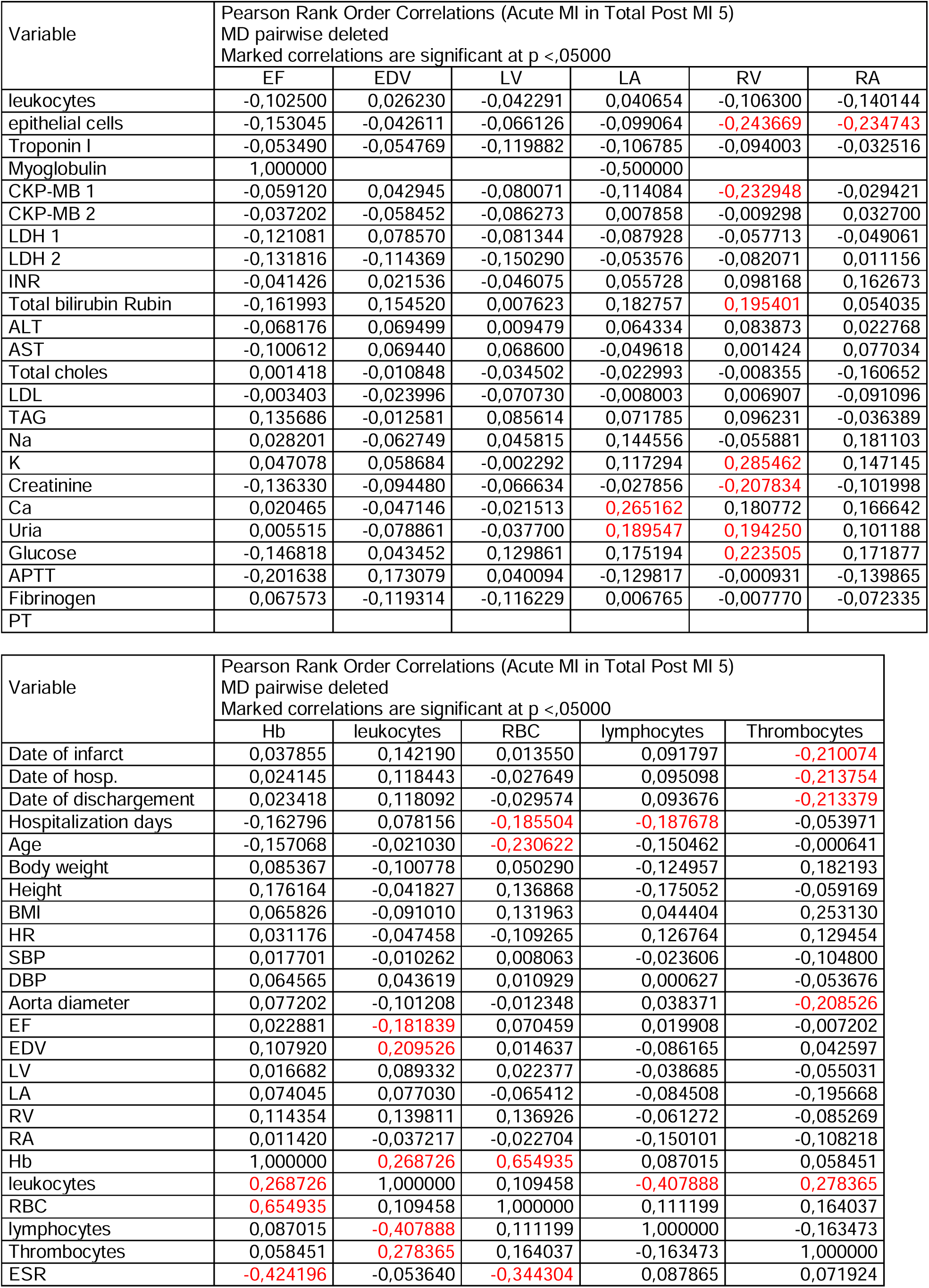
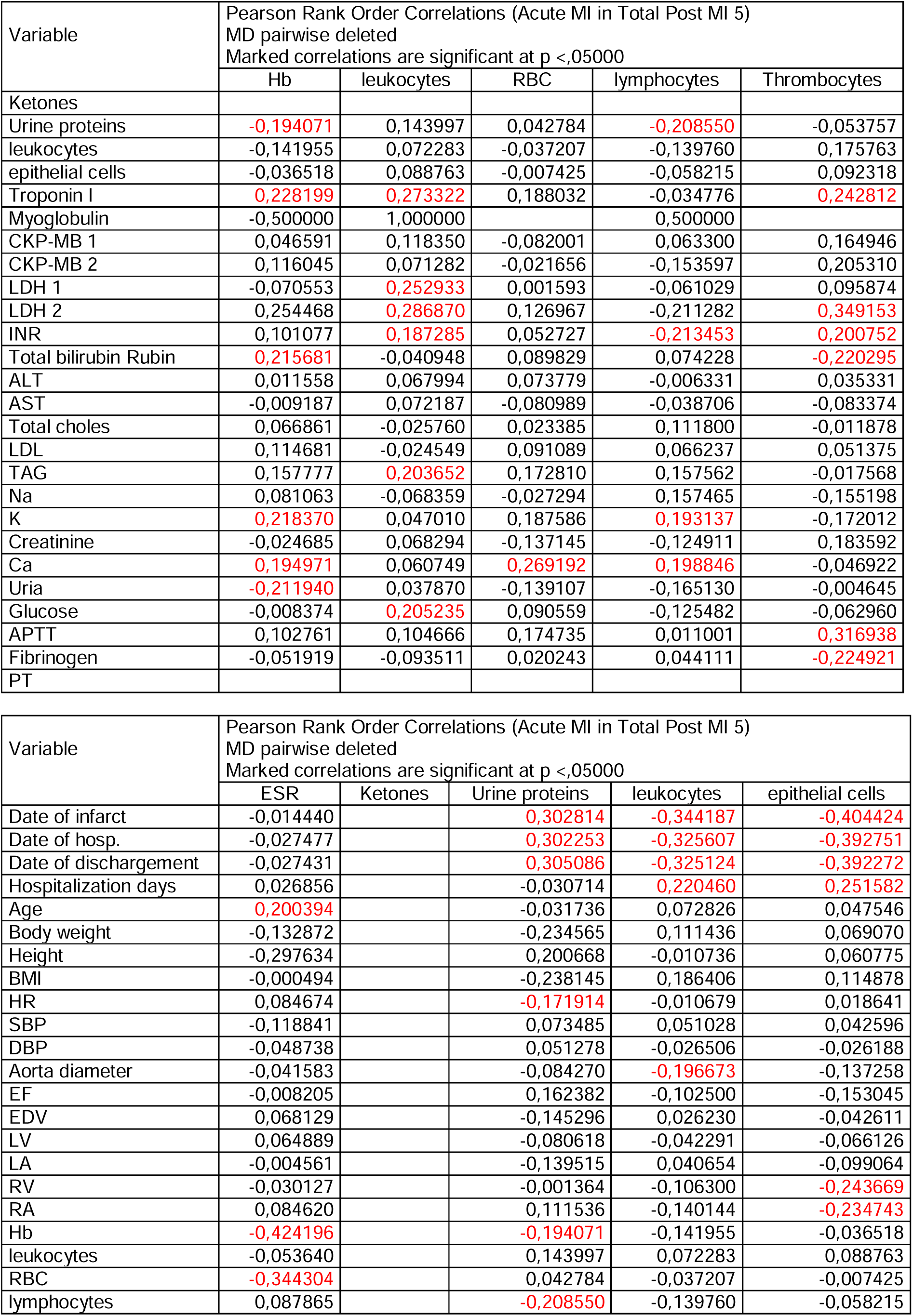
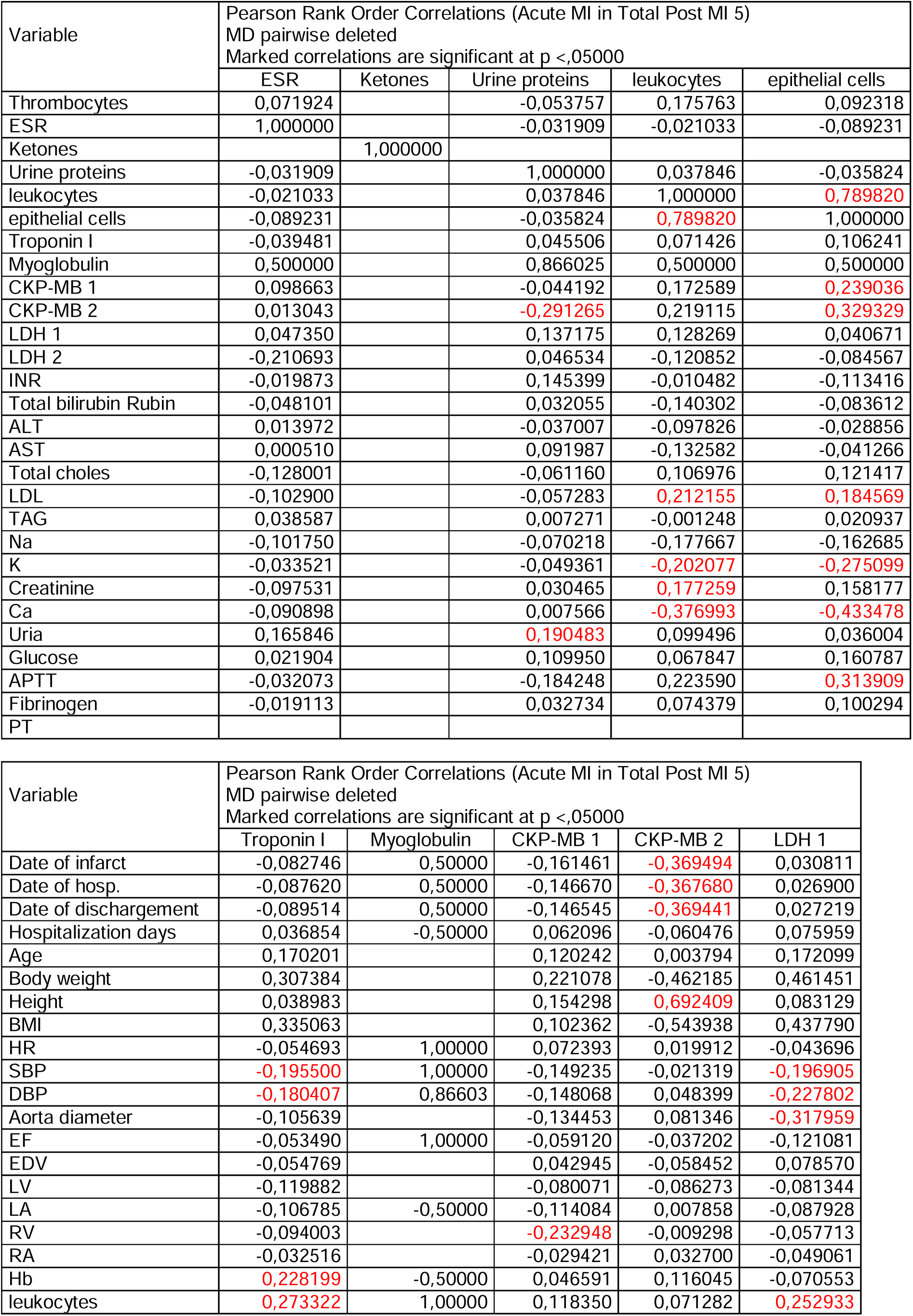
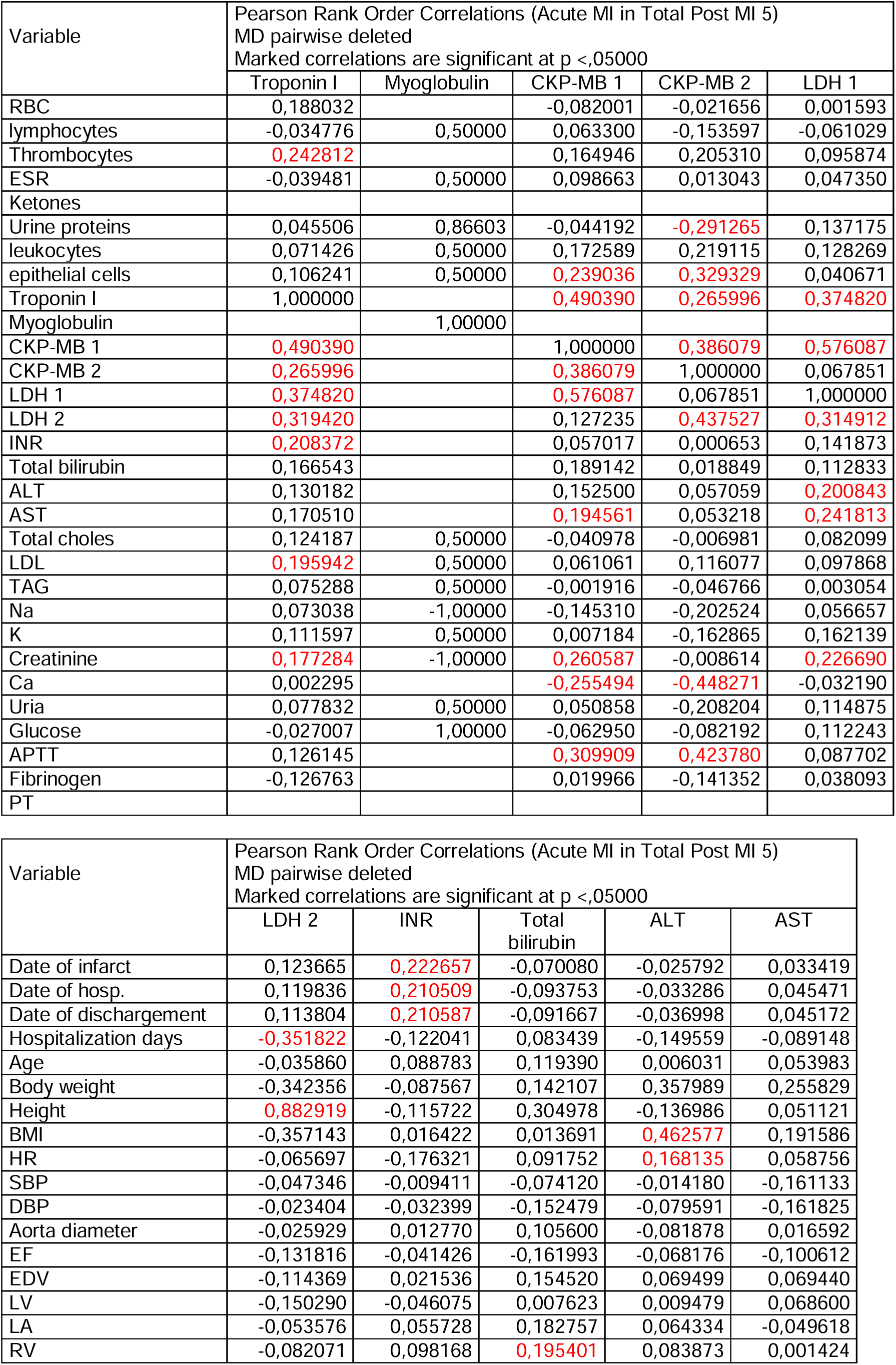
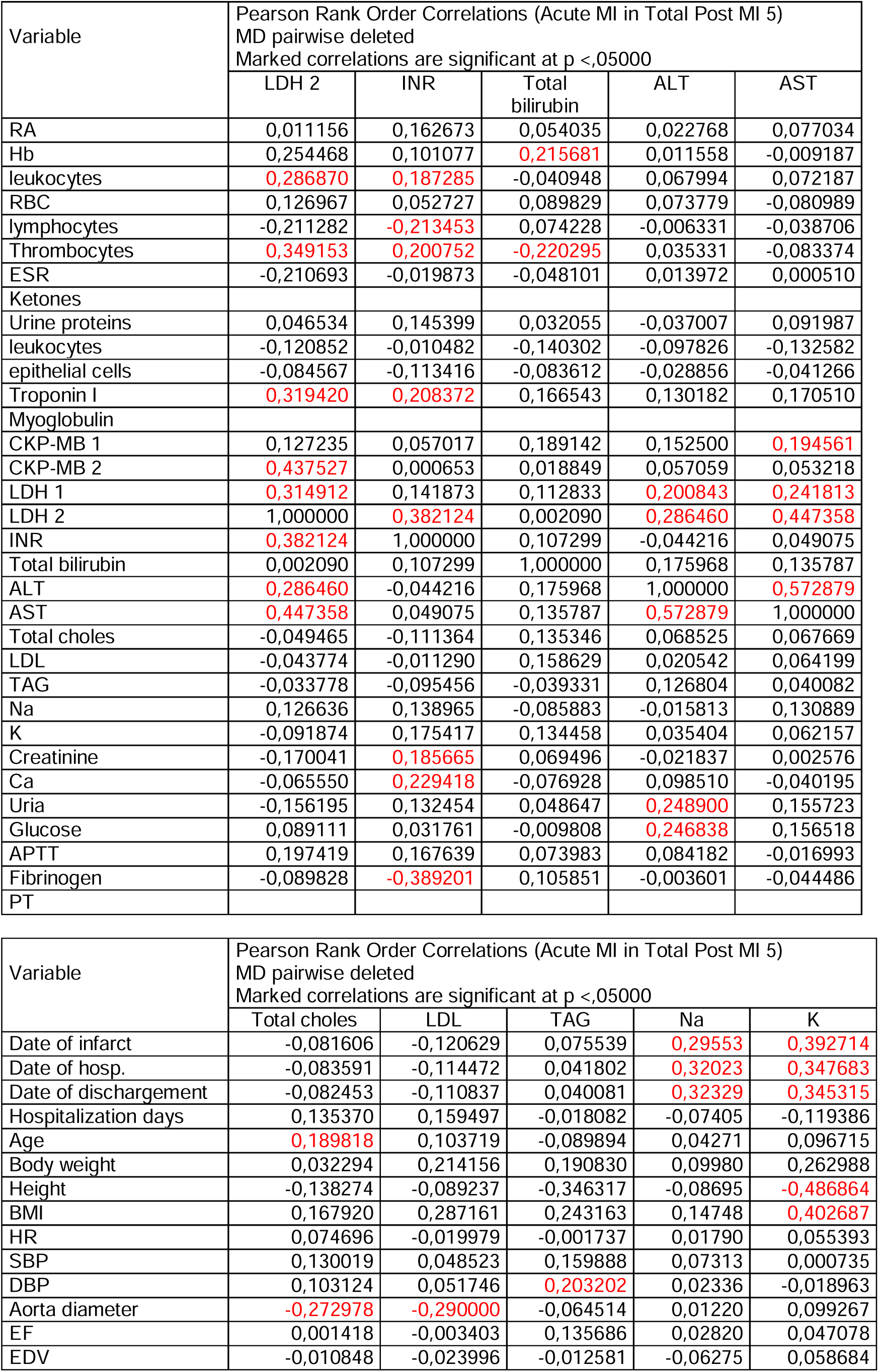
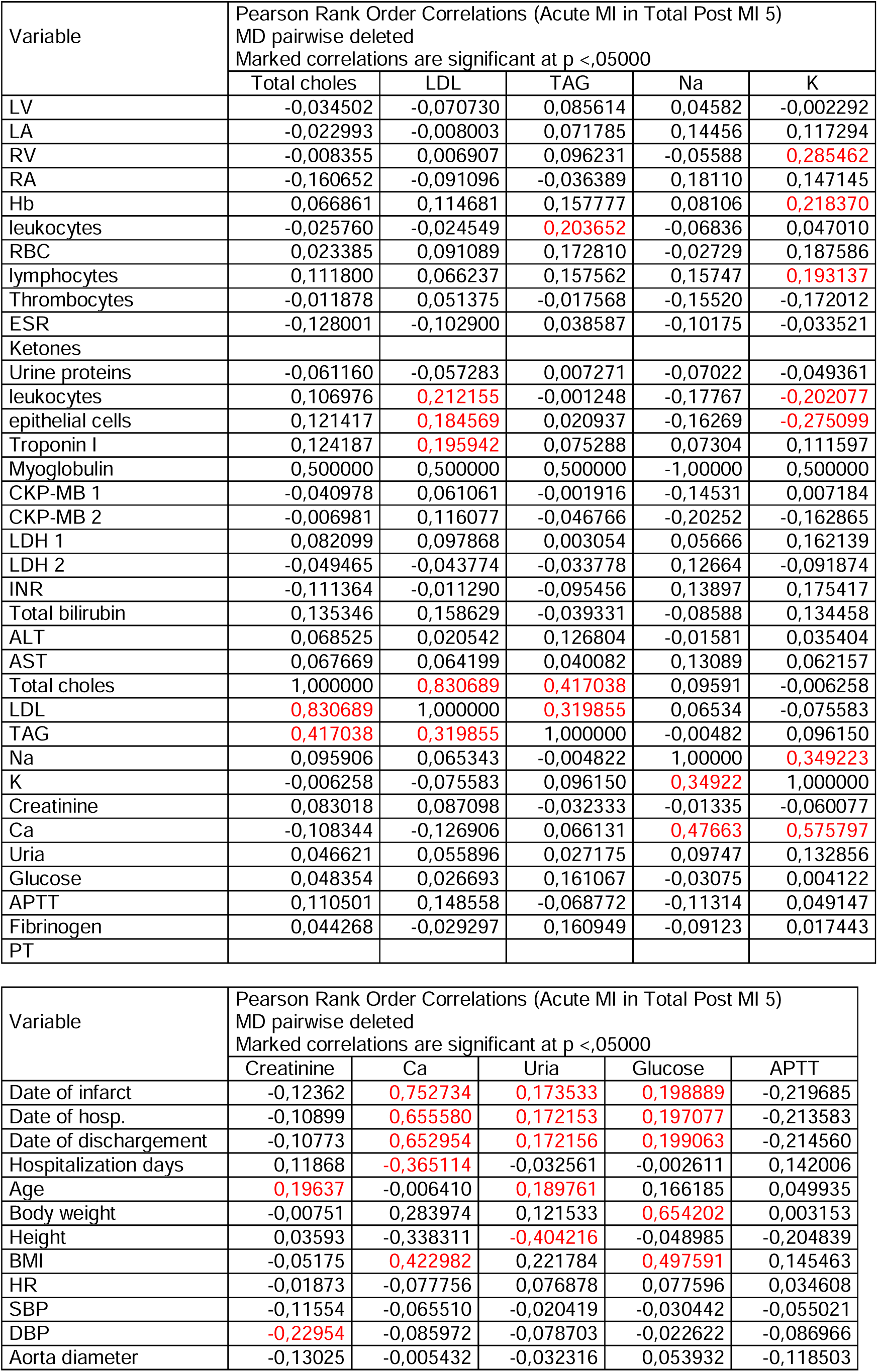
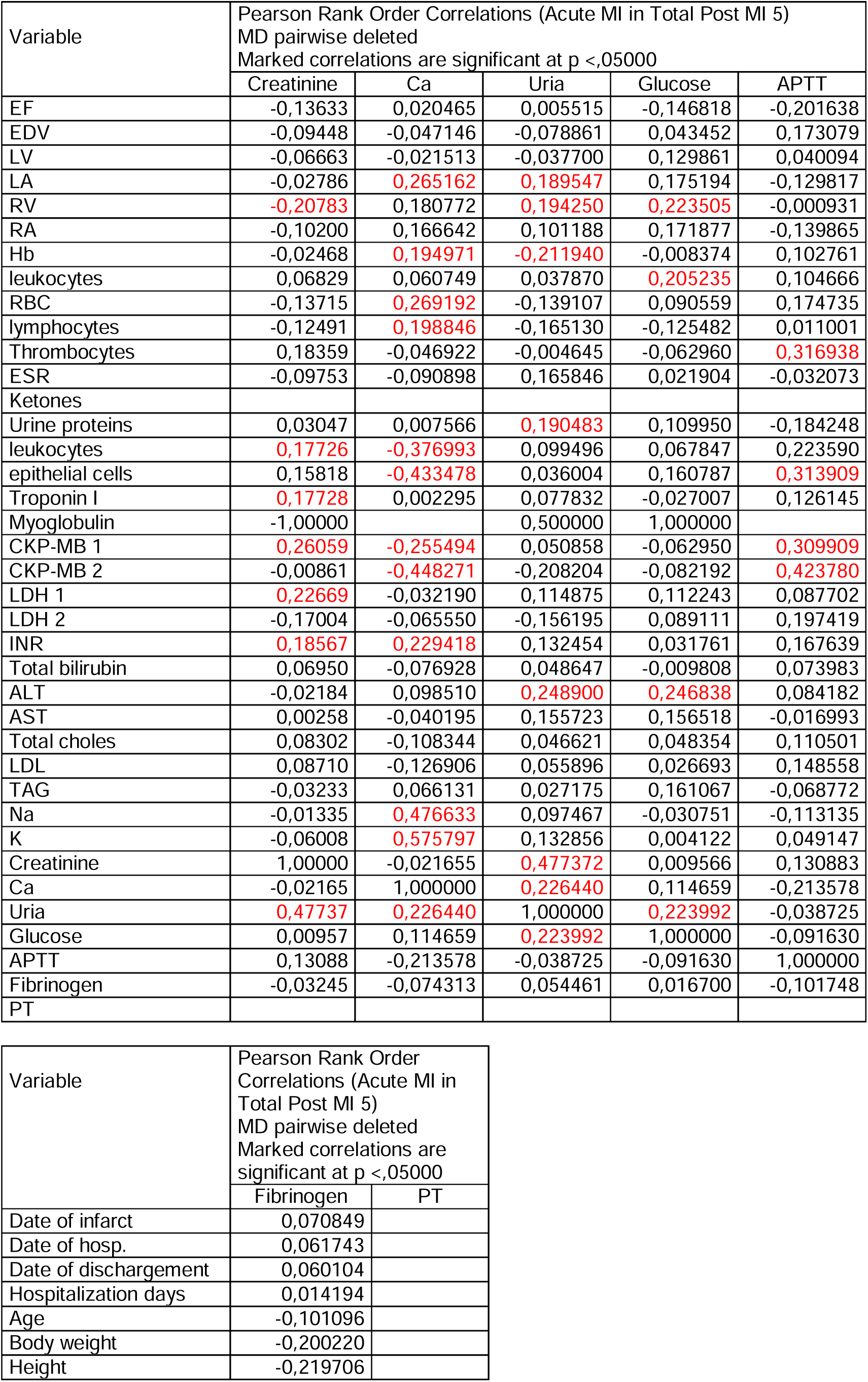
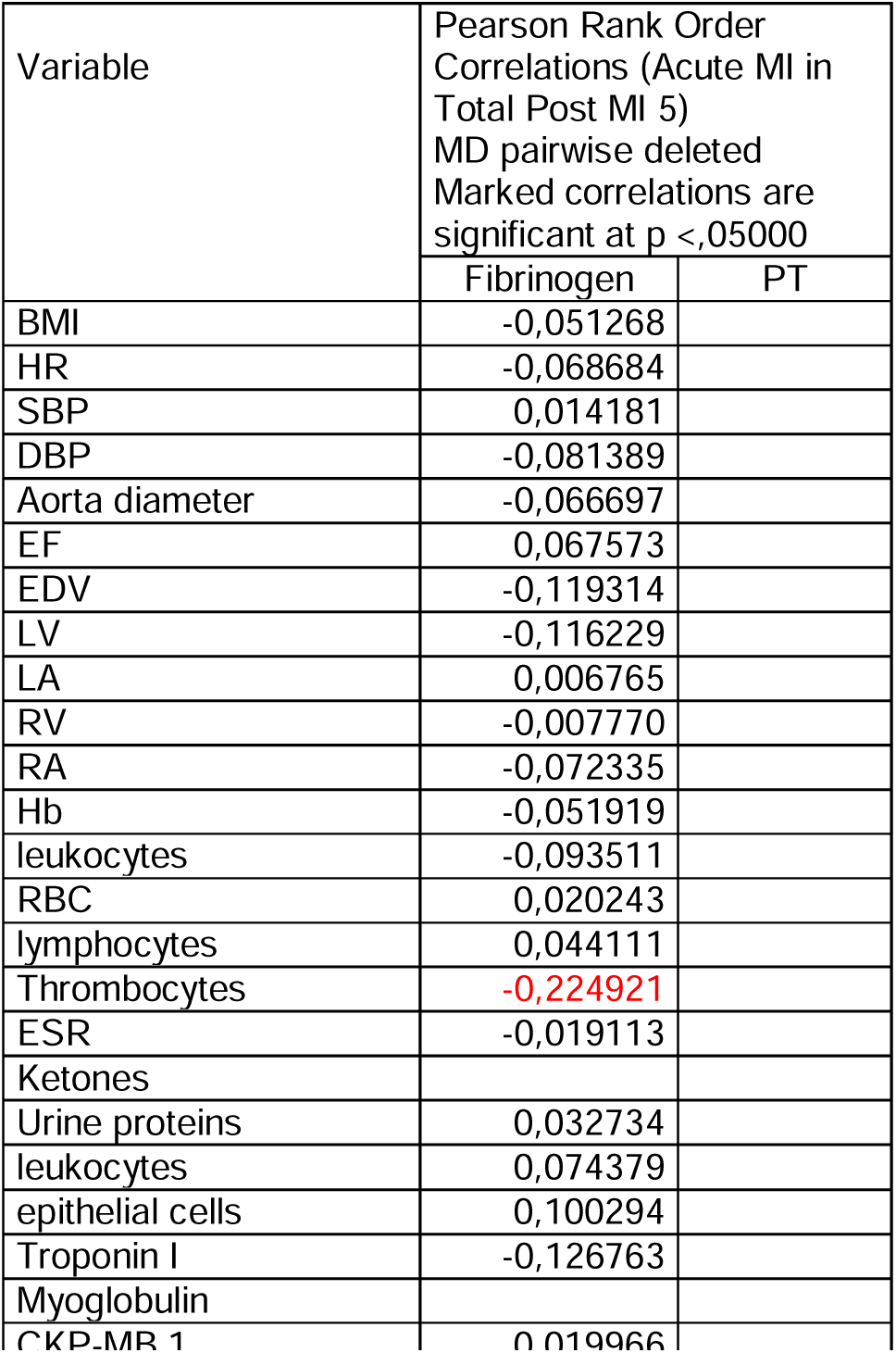
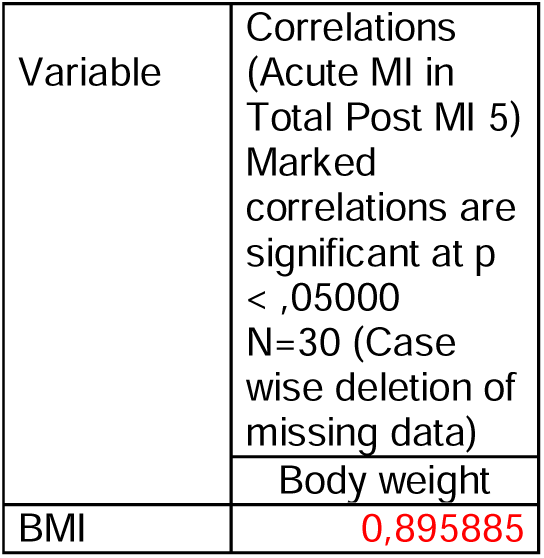

| Variable | Pearson Rank Order Correlations (Acute MI in Total Post MI 5)<br>MD pairwise deleted<br>Marked correlations are significant at $p < .05000$ | | |
| --- | --- | --- | --- |
|  | Date of infarct | Date of hosp. | Date of discharge |
| Date of infarct | 1,000000 | 0,998755 | 0,998662 |
| Date of hosp. | 0,998755 | 1,000000 | 0,999754 |
| Date of discharge | 0,998662 | 0,999754 | 1,000000 |
| Hospitalization days | -0,492819 | -0,480996 | -0,470104 |
| Age | -0,014406 | -0,017981 | -0,017534 |
| Body weight | -0,042648 | -0,056196 | -0,057004 |
| Height | -0,171595 | -0,175662 | -0,173637 |
| BMI | -0,050015 | -0,060710 | -0,063937 |
| HR | -0,026789 | -0,029367 | -0,028830 |
| SBP | 0,086786 | 0,096680 | 0,094712 |
| DBP | 0,050813 | 0,050794 | 0,050194 |
| Aorta diameter at the<br>Valsalva sinuses (mm) | -0,029993 | -0,022536 | -0,025733 |
| EF (%) | 0,080422 | 0,083670 | 0,083384 |
| EDV (ml) | -0,045009 | -0,050675 | -0,052638 |
| End diastolic LV size<br>(mm) | -0,001416 | -0,023427 | -0,022092 |
| End diastolic LA size<br>(mm) | 0,123265 | 0,117261 | 0,118687 |
| End diastolic RV size<br>(mm) | 0,224438 | 0,209813 | 0,208153 |
| End diastolic RA size<br>(mm) | 0,130204 | 0,133860 | 0,135790 |
| Hb | 0,037855 | 0,024145 | 0,023418 |
| leukocytes | 0,142190 | 0,118443 | 0,118092 |
| RBC | 0,013550 | -0,027649 | -0,029574 |
| lymphocytes | 0,091797 | 0,095098 | 0,093676 |
| Thrombocytes | -0,210074 | -0,213754 | -0,213379 |
| ESR | -0,014440 | -0,027477 | -0,027431 |
| Ketones |  |  |  |
| Urine proteins | 0,302814 | 0,302253 | 0,305086 |
| leukocytes | -0,344187 | -0,325607 | -0,325124 |
| epithelial cells | -0,404424 | -0,392751 | -0,392272 |
| Troponin I | -0,082746 | -0,087620 | -0,089514 |
| Myoglobin | 0,500000 | 0,500000 | 0,500000 |
| CKP-MB 1 | -0,161461 | -0,146670 | -0,146545 |
| CKP-MB 2 | -0,369494 | -0,367680 | -0,369441 |
| LDH 1 | 0,030811 | 0,026900 | 0,027219 |
| LDH 2 | 0,123665 | 0,119836 | 0,113804 |
| INR | 0,222657 | 0,210509 | 0,210587 |
| Total bilirubin | -0,070080 | -0,093753 | -0,091667 |
| ALT | -0,025792 | -0,033286 | -0,036998 |
| AST | 0,033419 | 0,045471 | 0,045172 |
| Total choles | -0,081606 | -0,083591 | -0,082453 |
| LDL | -0,120629 | -0,114472 | -0,110837 |
| TAG | 0,075539 | 0,041802 | 0,040081 |
| Na | 0,295534 | 0,320231 | 0,323290 |
| K | 0,392714 | 0,347683 | 0,345315 |
| Creatinine | -0,123623 | -0,108991 | -0,107730 |
| Ca | 0,752734 | 0,655580 | 0,652954 |
| Uria | 0,173533 | 0,172153 | 0,172156 |
|  | Date of infarct | Date of hosp. | Date of dischargement |
| Glucose | 0,198889 | 0,197077 | 0,199063 |
| APTT | -0,219685 | -0,213583 | -0,214560 |
| Fibrinogen | 0,070849 | 0,061743 | 0,060104 |
| PT |  |  |  |

| Variable | Pearson Rank Order Correlations (Acute MI in Total Post MI 5)<br>MD pairwise deleted<br>Marked correlations are significant at $p < .05000$ | | | |
| --- | --- | --- | --- | --- |
|  | Hospitalization days | Age | Body weight | Height |
| Date of infarct | -0,492819 | -0,014406 | -0,042648 | -0,171595 |
| Date of hosp. | -0,480996 | -0,017981 | -0,056196 | -0,175662 |
| Date of dischargement | -0,470104 | -0,017534 | -0,057004 | -0,173637 |
| Hospitalization days | 1,000000 | 0,092495 | -0,239856 | 0,084892 |
| Age | 0,092495 | 1,000000 | -0,056415 | 0,005208 |
| Body weight | -0,239856 | -0,056415 | 1,000000 | -0,178452 |
| Height | 0,084892 | 0,005208 | -0,178452 | 1,000000 |
| BMI | -0,336077 | 0,029871 | 0,898731 | -0,536541 |
| HR | 0,045764 | -0,060908 | 0,438935 | -0,249103 |
| SBP | -0,063265 | -0,054161 | 0,136715 | -0,045358 |
| DBP | 0,004338 | -0,110726 | 0,041841 | -0,011282 |
| Aorta diameter at the Valsalva sinuses (mm) | -0,056556 | -0,058935 | -0,108477 | 0,305970 |
| EF (%) | -0,041657 | -0,145918 | -0,120649 | -0,248037 |
| EDV (ml) | -0,066685 | -0,098928 | 0,438158 | 0,294385 |
| End diastolic LV size (mm) | 0,021873 | -0,076707 | 0,348287 | 0,273553 |
| End diastolic LA size (mm) | -0,006042 | 0,177610 | 0,177460 | 0,185164 |
| End diastolic RV size (mm) | -0,087129 | 0,037892 | 0,415737 | -0,152526 |
| End diastolic RA size (mm) | -0,018941 | 0,161934 | 0,180343 | 0,397011 |
| Hb | -0,162796 | -0,157068 | 0,085367 | 0,176164 |
| leukocytes | 0,078156 | -0,021030 | -0,100778 | -0,041827 |
| RBC | -0,185504 | -0,230622 | 0,050290 | 0,136868 |
| lymphocytes | -0,187678 | -0,150462 | -0,124957 | -0,175052 |
| Thrombocytes | -0,053971 | -0,000641 | 0,182193 | -0,059169 |
| ESR | 0,026856 | 0,200394 | -0,132872 | -0,297634 |
| Ketones |  |  |  |  |
| Urine proteins | -0,030714 | -0,031736 | -0,234565 | 0,200668 |
| leukocytes | 0,220460 | 0,072826 | 0,111436 | -0,010736 |
| epithelial cells | 0,251582 | 0,047546 | 0,069070 | 0,060775 |
| Troponin I | 0,036854 | 0,170201 | 0,307384 | 0,038983 |
| Myoglobin | -0,500000 |  |  |  |
| CKP-MB 1 | 0,062096 | 0,120242 | 0,221078 | 0,154298 |
| CKP-MB 2 | -0,060476 | 0,003794 | -0,462185 | 0,692409 |
| LDH 1 | 0,075959 | 0,172099 | 0,461451 | 0,083129 |
| LDH 2 | -0,351822 | -0,035860 | -0,342356 | 0,882919 |
| INR | -0,122041 | 0,088783 | -0,087567 | -0,115722 |
| Total bilirubin | 0,083439 | 0,119390 | 0,142107 | 0,304978 |
| ALT | -0,149559 | 0,006031 | 0,357989 | -0,136986 |

| Variable | Pearson Rank Order Correlations (Acute MI in Total Post MI 5)<br>MD pairwise deleted<br>Marked correlations are significant at p <,05000 |  |  |  |
| --- | --- | --- | --- | --- |
|  | Hospitalization days | Age | Body weight | Height |
| AST | -0,089148 | 0,053983 | 0,255829 | 0,051121 |
| Total choles | 0,135370 | 0,189818 | 0,032294 | -0,138274 |
| LDL | 0,159497 | 0,103719 | 0,214156 | -0,089237 |
| TAG | -0,018082 | -0,089894 | 0,190830 | -0,346317 |
| Na | -0,074054 | 0,042710 | 0,099801 | -0,086951 |
| K | -0,119386 | 0,096715 | 0,262988 | -0,486864 |
| Creatinine | 0,118681 | 0,196371 | -0,007508 | 0,035929 |
| Ca | -0,365114 | -0,006410 | 0,283974 | -0,338311 |
| Uria | -0,032561 | 0,189761 | 0,121533 | -0,404216 |
| Glucose | -0,002611 | 0,166185 | 0,654202 | -0,048985 |
| APTT | 0,142006 | 0,049935 | 0,003153 | -0,204839 |
| Fibrinogen | 0,014194 | -0,101096 | -0,200220 | -0,219706 |
| PT |  |  |  |  |

| Variable | Pearson Rank Order Correlations (Acute MI in Total Post MI 5)<br>MD pairwise deleted<br>Marked correlations are significant at p <,05000 |  |  |  |  |
| --- | --- | --- | --- | --- | --- |
|  | BMI | HR | SBP | DBP | Aorta diameter |
| Date of infarct | -0,050015 | -0,026789 | 0,086786 | 0,050813 | -0,029993 |
| Date of hosp. | -0,060710 | -0,029367 | 0,096680 | 0,050794 | -0,022536 |
| Date of dischargement | -0,063937 | -0,028830 | 0,094712 | 0,050194 | -0,025733 |
| Hospitalization days | -0,336077 | 0,045764 | -0,063265 | 0,004338 | -0,056556 |
| Age | 0,029871 | -0,060908 | -0,054161 | -0,110726 | -0,058935 |
| Body weight | 0,898731 | 0,438935 | 0,136715 | 0,041841 | -0,108477 |
| Height | -0,536541 | -0,249103 | -0,045358 | -0,011282 | 0,305970 |
| BMI | 1,000000 | 0,475827 | 0,233983 | 0,134203 | -0,303926 |
| HR | 0,475827 | 1,000000 | 0,164042 | 0,193671 | 0,056941 |
| SBP | 0,233983 | 0,164042 | 1,000000 | 0,837480 | 0,062520 |
| DBP | 0,134203 | 0,193671 | 0,837480 | 1,000000 | 0,109910 |
| Aorta diameter at the Valsalva sinuses (mm) | -0,303926 | 0,056941 | 0,062520 | 0,109910 | 1,000000 |
| EF (%) | -0,032926 | -0,178774 | 0,093906 | 0,042189 | 0,050386 |
| EDV (ml) | 0,321078 | 0,012821 | 0,129426 | 0,181506 | 0,126137 |
| End diastolic LV size (mm) | 0,062654 | 0,108568 | 0,078351 | 0,214669 | 0,217956 |
| End diastolic LA size (mm) | -0,090691 | -0,033390 | 0,127778 | 0,117089 | 0,220792 |
| End diastolic RV size (mm) | 0,343756 | 0,122876 | 0,107076 | 0,187414 | 0,339438 |
| End diastolic RA size (mm) | -0,040683 | 0,009769 | -0,000967 | 0,056850 | 0,222980 |
| Hb | 0,065826 | 0,031176 | 0,017701 | 0,064565 | 0,077202 |
| leukocytes | -0,091010 | -0,047458 | -0,010262 | 0,043619 | -0,101208 |
| RBC | 0,131963 | -0,109265 | 0,008063 | 0,010929 | -0,012348 |
| lymphocytes | 0,044404 | 0,126764 | -0,023606 | 0,000627 | 0,038371 |
| Thrombocytes | 0,253130 | 0,129454 | -0,104800 | -0,053676 | -0,208526 |
| ESR | -0,000494 | 0,084674 | -0,118841 | -0,048738 | -0,041583 |
| Ketones |  |  |  |  |  |
| Urine proteins | -0,238145 | -0,171914 | 0,073485 | 0,051278 | -0,084270 |
| leukocytes | 0,186406 | -0,010679 | 0,051028 | -0,026506 | -0,196673 |
| epithelial cells | 0,114878 | 0,018641 | 0,042596 | -0,026188 | -0,137258 |

| Variable | Pearson Rank Order Correlations (Acute MI in Total Post MI 5) |  |  |  |  |
| --- | --- | --- | --- | --- | --- |
| | MD pairwise deleted<br>Marked correlations are significant at $p < .05000$ | | | | |
|  | BMI | HR | SBP | DBP | Aorta diameter |
| Troponin I | 0,335063 | -0,054693 | -0,195500 | -0,180407 | -0,105639 |
| Myoglobin |  | 1,000000 | 1,000000 | 0,866025 |  |
| CKP-MB 1 | 0,102362 | 0,072393 | -0,149235 | -0,148068 | -0,134453 |
| CKP-MB 2 | -0,543938 | 0,019912 | -0,021319 | 0,048399 | 0,081346 |
| LDH 1 | 0,437790 | -0,043696 | -0,196905 | -0,227802 | -0,317959 |
| LDH 2 | -0,357143 | -0,065697 | -0,047346 | -0,023404 | -0,025929 |
| INR | 0,016422 | -0,176321 | -0,009411 | -0,032399 | 0,012770 |
| Total bilirubin Rubin | 0,013691 | 0,091752 | -0,074120 | -0,152479 | 0,105600 |
| ALT | 0,462577 | 0,168135 | -0,014180 | -0,079591 | -0,081878 |
| AST | 0,191586 | 0,058756 | -0,161133 | -0,161825 | 0,016592 |
| Total choles | 0,167920 | 0,074696 | 0,130019 | 0,103124 | -0,272978 |
| LDL | 0,287161 | -0,019979 | 0,048523 | 0,051746 | -0,290000 |
| TAG | 0,243163 | -0,001737 | 0,159888 | 0,203202 | -0,064514 |
| Na | 0,147481 | 0,017900 | 0,073128 | 0,023359 | 0,012202 |
| K | 0,402687 | 0,055393 | 0,000735 | -0,018963 | 0,099267 |
| Creatinine | -0,051752 | -0,018731 | -0,115542 | -0,229538 | -0,130249 |
| Ca | 0,422982 | -0,077756 | -0,065510 | -0,085972 | -0,005432 |
| Uria | 0,221784 | 0,076878 | -0,020419 | -0,078703 | -0,032316 |
| Glucose | 0,497591 | 0,077596 | -0,030442 | -0,022622 | 0,053932 |
| APTT | 0,145463 | 0,034608 | -0,055021 | -0,086966 | -0,118503 |
| Fibrinogen | -0,051268 | -0,068684 | 0,014181 | -0,081389 | -0,066697 |
| PT |  |  |  |  |  |

| Variable | Pearson Rank Order Correlations (Acute MI in Total Post MI 5) |  |  |  |  |  |
| --- | --- | --- | --- | --- | --- | --- |
| | MD pairwise deleted<br>Marked correlations are significant at $p < .05000$ | | | | | |
|  | EF | EDV | LV | LA | RV | RA |
| Date of infarct | 0,080422 | -0,045009 | -0,001416 | 0,123265 | 0,224438 | 0,130204 |
| Date of hosp. | 0,083670 | -0,050675 | -0,023427 | 0,117261 | 0,209813 | 0,133860 |
| Date of dischargement | 0,083384 | -0,052638 | -0,022092 | 0,118687 | 0,208153 | 0,135790 |
| Hospitalization days | -0,041657 | -0,066685 | 0,021873 | -0,006042 | -0,087129 | -0,018941 |
| Age | -0,145918 | -0,098928 | -0,076707 | 0,177610 | 0,037892 | 0,161934 |
| Body weight | -0,120649 | 0,438158 | 0,348287 | 0,177460 | 0,415737 | 0,180343 |
| Height | -0,248037 | 0,294385 | 0,273553 | 0,185164 | -0,152526 | 0,397011 |
| BMI | -0,032926 | 0,321078 | 0,062654 | -0,090691 | 0,343756 | -0,040683 |
| HR | -0,178774 | 0,012821 | 0,108568 | -0,033390 | 0,122876 | 0,009769 |
| SBP | 0,093906 | 0,129426 | 0,078351 | 0,127778 | 0,107076 | -0,000967 |
| DBP | 0,042189 | 0,181506 | 0,214669 | 0,117089 | 0,187414 | 0,056850 |
| Aorta diameter | 0,050386 | 0,126137 | 0,217956 | 0,220792 | 0,339438 | 0,222980 |
| EF | 1,000000 | -0,204007 | -0,306233 | -0,037623 | 0,037063 | 0,051806 |
| EDV | -0,204007 | 1,000000 | 0,816552 | 0,427731 | 0,232208 | 0,153922 |
| LV | -0,306233 | 0,816552 | 1,000000 | 0,373137 | 0,198793 | 0,180908 |
| LA | -0,037623 | 0,427731 | 0,373137 | 1,000000 | 0,421956 | 0,553528 |
| RV | 0,037063 | 0,232208 | 0,198793 | 0,421956 | 1,000000 | 0,222865 |
| RA | 0,051806 | 0,153922 | 0,180908 | 0,553528 | 0,222865 | 1,000000 |
| Hb | 0,022881 | 0,107920 | 0,016682 | 0,074045 | 0,114354 | 0,011420 |
| leukocytes | -0,181839 | 0,209526 | 0,089332 | 0,077030 | 0,139811 | -0,037217 |
| RBC | 0,070459 | 0,014637 | 0,022377 | -0,065412 | 0,136926 | -0,022704 |
| lymphocytes | 0,019908 | -0,086165 | -0,038685 | -0,084508 | -0,061272 | -0,150101 |
| Thrombocytes | -0,007202 | 0,042597 | -0,055031 | -0,195668 | -0,085269 | -0,108218 |
| ESR | -0,008205 | 0,068129 | 0,064889 | -0,004561 | -0,030127 | 0,084620 |
| Ketones |  |  |  |  |  |  |
| Urine proteins | 0,162382 | -0,145296 | -0,080618 | -0,139515 | -0,001364 | 0,111536 |
| | MD pairwise deleted<br>Marked correlations are significant at $p < .05000$ | | | | | |
|  | EF | EDV | LV | LA | RV | RA |
| leukocytes | -0,102500 | 0,026230 | -0,042291 | 0,040654 | -0,106300 | -0,140144 |
| epithelial cells | -0,153045 | -0,042611 | -0,066126 | -0,099064 | -0,243669 | -0,234743 |
| Troponin I | -0,053490 | -0,054769 | -0,119882 | -0,106785 | -0,094003 | -0,032516 |
| Myoglobin | 1,000000 |  |  | -0,500000 |  |  |
| CKP-MB 1 | -0,059120 | 0,042945 | -0,080071 | -0,114084 | -0,232948 | -0,029421 |
| CKP-MB 2 | -0,037202 | -0,058452 | -0,086273 | 0,007858 | -0,009298 | 0,032700 |
| LDH 1 | -0,121081 | 0,078570 | -0,081344 | -0,087928 | -0,057713 | -0,049061 |
| LDH 2 | -0,131816 | -0,114369 | -0,150290 | -0,053576 | -0,082071 | 0,011156 |
| INR | -0,041426 | 0,021536 | -0,046075 | 0,055728 | 0,098168 | 0,162673 |
| Total bilirubin Rubin | -0,161993 | 0,154520 | 0,007623 | 0,182757 | 0,195401 | 0,054035 |
| ALT | -0,068176 | 0,069499 | 0,009479 | 0,064334 | 0,083873 | 0,022768 |
| AST | -0,100612 | 0,069440 | 0,068600 | -0,049618 | 0,001424 | 0,077034 |
| Total choles | 0,001418 | -0,010848 | -0,034502 | -0,022993 | -0,008355 | -0,160652 |
| LDL | -0,003403 | -0,023996 | -0,070730 | -0,008003 | 0,006907 | -0,091096 |
| TAG | 0,135686 | -0,012581 | 0,085614 | 0,071785 | 0,096231 | -0,036389 |
| Na | 0,028201 | -0,062749 | 0,045815 | 0,144556 | -0,055881 | 0,181103 |
| K | 0,047078 | 0,058684 | -0,002292 | 0,117294 | 0,285462 | 0,147145 |
| Creatinine | -0,136330 | -0,094480 | -0,066634 | -0,027856 | -0,207834 | -0,101998 |
| Ca | 0,020465 | -0,047146 | -0,021513 | 0,265162 | 0,180772 | 0,166642 |
| Uria | 0,005515 | -0,078861 | -0,037700 | 0,189547 | 0,194250 | 0,101188 |
| Glucose | -0,146818 | 0,043452 | 0,129861 | 0,175194 | 0,223505 | 0,171877 |
| APTT | -0,201638 | 0,173079 | 0,040094 | -0,129817 | -0,000931 | -0,139865 |
| Fibrinogen | 0,067573 | -0,119314 | -0,116229 | 0,006765 | -0,007770 | -0,072335 |
| PT |  |  |  |  |  |  |

| Variable | Pearson Rank Order Correlations (Acute MI in Total Post MI 5) |  |  |  |  |
| --- | --- | --- | --- | --- | --- |
| | MD pairwise deleted<br>Marked correlations are significant at $p < .05000$ | | | | |
|  | Hb | leukocytes | RBC | lymphocytes | Thrombocytes |
| Date of infarct | 0,037855 | 0,142190 | 0,013550 | 0,091797 | -0,210074 |
| Date of hosp. | 0,024145 | 0,118443 | -0,027649 | 0,095098 | -0,213754 |
| Date of dischargement | 0,023418 | 0,118092 | -0,029574 | 0,093676 | -0,213379 |
| Hospitalization days | -0,162796 | 0,078156 | -0,185504 | -0,187678 | -0,053971 |
| Age | -0,157068 | -0,021030 | -0,230622 | -0,150462 | -0,000641 |
| Body weight | 0,085367 | -0,100778 | 0,050290 | -0,124957 | 0,182193 |
| Height | 0,176164 | -0,041827 | 0,136868 | -0,175052 | -0,059169 |
| BMI | 0,065826 | -0,091010 | 0,131963 | 0,044404 | 0,253130 |
| HR | 0,031176 | -0,047458 | -0,109265 | 0,126764 | 0,129454 |
| SBP | 0,017701 | -0,010262 | 0,008063 | -0,023606 | -0,104800 |
| DBP | 0,064565 | 0,043619 | 0,010929 | 0,000627 | -0,053676 |
| Aorta diameter | 0,077202 | -0,101208 | -0,012348 | 0,038371 | -0,208526 |
| EF | 0,022881 | -0,181839 | 0,070459 | 0,019908 | -0,007202 |
| EDV | 0,107920 | 0,209526 | 0,014637 | -0,086165 | 0,042597 |
| LV | 0,016682 | 0,089332 | 0,022377 | -0,038685 | -0,055031 |
| LA | 0,074045 | 0,077030 | -0,065412 | -0,084508 | -0,195668 |
| RV | 0,114354 | 0,139811 | 0,136926 | -0,061272 | -0,085269 |
| RA | 0,011420 | -0,037217 | -0,022704 | -0,150101 | -0,108218 |
| Hb | 1,000000 | 0,268726 | 0,654935 | 0,087015 | 0,058451 |
| leukocytes | 0,268726 | 1,000000 | 0,109458 | -0,407888 | 0,278365 |
| RBC | 0,654935 | 0,109458 | 1,000000 | 0,111199 | 0,164037 |
| lymphocytes | 0,087015 | -0,407888 | 0,111199 | 1,000000 | -0,163473 |
| Thrombocytes | 0,058451 | 0,278365 | 0,164037 | -0,163473 | 1,000000 |
| ESR | -0,424196 | -0,053640 | -0,344304 | 0,087865 | 0,071924 |

| Variable | Pearson Rank Order Correlations (Acute MI in Total Post MI 5)<br>MD pairwise deleted<br>Marked correlations are significant at p <,05000 |  |  |  |  |
| --- | --- | --- | --- | --- | --- |
|  | Hb | leukocytes | RBC | lymphocytes | Thrombocytes |
| Ketones |  |  |  |  |  |
| Urine proteins | -0,194071 | 0,143997 | 0,042784 | -0,208550 | -0,053757 |
| leukocytes | -0,141955 | 0,072283 | -0,037207 | -0,139760 | 0,175763 |
| epithelial cells | -0,036518 | 0,088763 | -0,007425 | -0,058215 | 0,092318 |
| Troponin I | 0,228199 | 0,273322 | 0,188032 | -0,034776 | 0,242812 |
| Myoglobin | -0,500000 | 1,000000 |  | 0,500000 |  |
| CKP-MB 1 | 0,046591 | 0,118350 | -0,082001 | 0,063300 | 0,164946 |
| CKP-MB 2 | 0,116045 | 0,071282 | -0,021656 | -0,153597 | 0,205310 |
| LDH 1 | -0,070553 | 0,252933 | 0,001593 | -0,061029 | 0,095874 |
| LDH 2 | 0,254468 | 0,286870 | 0,126967 | -0,211282 | 0,349153 |
| INR | 0,101077 | 0,187285 | 0,052727 | -0,213453 | 0,200752 |
| Total bilirubin Rubin | 0,215681 | -0,040948 | 0,089829 | 0,074228 | -0,220295 |
| ALT | 0,011558 | 0,067994 | 0,073779 | -0,006331 | 0,035331 |
| AST | -0,009187 | 0,072187 | -0,080989 | -0,038706 | -0,083374 |
| Total choles | 0,066861 | -0,025760 | 0,023385 | 0,111800 | -0,011878 |
| LDL | 0,114681 | -0,024549 | 0,091089 | 0,066237 | 0,051375 |
| TAG | 0,157777 | 0,203652 | 0,172810 | 0,157562 | -0,017568 |
| Na | 0,081063 | -0,068359 | -0,027294 | 0,157465 | -0,155198 |
| K | 0,218370 | 0,047010 | 0,187586 | 0,193137 | -0,172012 |
| Creatinine | -0,024685 | 0,068294 | -0,137145 | -0,124911 | 0,183592 |
| Ca | 0,194971 | 0,060749 | 0,269192 | 0,198846 | -0,046922 |
| Uria | -0,211940 | 0,037870 | -0,139107 | -0,165130 | -0,004645 |
| Glucose | -0,008374 | 0,205235 | 0,090559 | -0,125482 | -0,062960 |
| APTT | 0,102761 | 0,104666 | 0,174735 | 0,011001 | 0,316938 |
| Fibrinogen | -0,051919 | -0,093511 | 0,020243 | 0,044111 | -0,224921 |
| PT |  |  |  |  |  |
|  | ESR | Ketones | Urine proteins | leukocytes | epithelial cells |
| Date of infarct | -0,014440 |  | 0,302814 | -0,344187 | -0,404424 |
| Date of hosp. | -0,027477 |  | 0,302253 | -0,325607 | -0,392751 |
| Date of dischargement | -0,027431 |  | 0,305086 | -0,325124 | -0,392272 |
| Hospitalization days | 0,026856 |  | -0,030714 | 0,220460 | 0,251582 |
| Age | 0,200394 |  | -0,031736 | 0,072826 | 0,047546 |
| Body weight | -0,132872 |  | -0,234565 | 0,111436 | 0,069070 |
| Height | -0,297634 |  | 0,200668 | -0,010736 | 0,060775 |
| BMI | -0,000494 |  | -0,238145 | 0,186406 | 0,114878 |
| HR | 0,084674 |  | -0,171914 | -0,010679 | 0,018641 |
| SBP | -0,118841 |  | 0,073485 | 0,051028 | 0,042596 |
| DBP | -0,048738 |  | 0,051278 | -0,026506 | -0,026188 |
| Aorta diameter | -0,041583 |  | -0,084270 | -0,196673 | -0,137258 |
| EF | -0,008205 |  | 0,162382 | -0,102500 | -0,153045 |
| EDV | 0,068129 |  | -0,145296 | 0,026230 | -0,042611 |
| LV | 0,064889 |  | -0,080618 | -0,042291 | -0,066126 |
| LA | -0,004561 |  | -0,139515 | 0,040654 | -0,099064 |
| RV | -0,030127 |  | -0,001364 | -0,106300 | -0,243669 |
| RA | 0,084620 |  | 0,111536 | -0,140144 | -0,234743 |
| Hb | -0,424196 |  | -0,194071 | -0,141955 | -0,036518 |
| leukocytes | -0,053640 |  | 0,143997 | 0,072283 | 0,088763 |
| RBC | -0,344304 |  | 0,042784 | -0,037207 | -0,007425 |
| lymphocytes | 0,087865 |  | -0,208550 | -0,139760 | -0,058215 |
|  | ESR | Ketones | Urine proteins | leukocytes | epithelial cells |
| Thrombocytes | 0,071924 |  | -0,053757 | 0,175763 | 0,092318 |
| ESR | 1,000000 |  | -0,031909 | -0,021033 | -0,089231 |
| Ketones |  | 1,000000 |  |  |  |
| Urine proteins | -0,031909 |  | 1,000000 | 0,037846 | -0,035824 |
| leukocytes | -0,021033 |  | 0,037846 | 1,000000 | 0,789820 |
| epithelial cells | -0,089231 |  | -0,035824 | 0,789820 | 1,000000 |
| Troponin I | -0,039481 |  | 0,045506 | 0,071426 | 0,106241 |
| Myoglobulin | 0,500000 |  | 0,866025 | 0,500000 | 0,500000 |
| CKP-MB 1 | 0,098663 |  | -0,044192 | 0,172589 | 0,239036 |
| CKP-MB 2 | 0,013043 |  | -0,291265 | 0,219115 | 0,329329 |
| LDH 1 | 0,047350 |  | 0,137175 | 0,128269 | 0,040671 |
| LDH 2 | -0,210693 |  | 0,046534 | -0,120852 | -0,084567 |
| INR | -0,019873 |  | 0,145399 | -0,010482 | -0,113416 |
| Total bilirubin Rubin | -0,048101 |  | 0,032055 | -0,140302 | -0,083612 |
| ALT | 0,013972 |  | -0,037007 | -0,097826 | -0,028856 |
| AST | 0,000510 |  | 0,091987 | -0,132582 | -0,041266 |
| Total choles | -0,128001 |  | -0,061160 | 0,106976 | 0,121417 |
| LDL | -0,102900 |  | -0,057283 | 0,212155 | 0,184569 |
| TAG | 0,038587 |  | 0,007271 | -0,001248 | 0,020937 |
| Na | -0,101750 |  | -0,070218 | -0,177667 | -0,162685 |
| K | -0,033521 |  | -0,049361 | -0,202077 | -0,275099 |
| Creatinine | -0,097531 |  | 0,030465 | 0,177259 | 0,158177 |
| Ca | -0,090898 |  | 0,007566 | -0,376993 | -0,433478 |
| Uria | 0,165846 |  | 0,190483 | 0,099496 | 0,036004 |
| Glucose | 0,021904 |  | 0,109950 | 0,067847 | 0,160787 |
| APTT | -0,032073 |  | -0,184248 | 0,223590 | 0,313909 |
| Fibrinogen | -0,019113 |  | 0,032734 | 0,074379 | 0,100294 |
| PT |  |  |  |  |  |
|  | Troponin I | Myoglobulin | CKP-MB 1 | CKP-MB 2 | LDH 1 |
| Date of infarct | -0,082746 | 0,50000 | -0,161461 | -0,369494 | 0,030811 |
| Date of hosp. | -0,087620 | 0,50000 | -0,146670 | -0,367680 | 0,026900 |
| Date of dischargement | -0,089514 | 0,50000 | -0,146545 | -0,369441 | 0,027219 |
| Hospitalization days | 0,036854 | -0,50000 | 0,062096 | -0,060476 | 0,075959 |
| Age | 0,170201 |  | 0,120242 | 0,003794 | 0,172099 |
| Body weight | 0,307384 |  | 0,221078 | -0,462185 | 0,461451 |
| Height | 0,038983 |  | 0,154298 | 0,692409 | 0,083129 |
| BMI | 0,335063 |  | 0,102362 | -0,543938 | 0,437790 |
| HR | -0,054693 | 1,00000 | 0,072393 | 0,019912 | -0,043696 |
| SBP | -0,195500 | 1,00000 | -0,149235 | -0,021319 | -0,196905 |
| DBP | -0,180407 | 0,86603 | -0,148068 | 0,048399 | -0,227802 |
| Aorta diameter | -0,105639 |  | -0,134453 | 0,081346 | -0,317959 |
| EF | -0,053490 | 1,00000 | -0,059120 | -0,037202 | -0,121081 |
| EDV | -0,054769 |  | 0,042945 | -0,058452 | 0,078570 |
| LV | -0,119882 |  | -0,080071 | -0,086273 | -0,081344 |
| LA | -0,106785 | -0,50000 | -0,114084 | 0,007858 | -0,087928 |
| RV | -0,094003 |  | -0,232948 | -0,009298 | -0,057713 |
| RA | -0,032516 |  | -0,029421 | 0,032700 | -0,049061 |
| Hb | 0,228199 | -0,50000 | 0,046591 | 0,116045 | -0,070553 |
| leukocytes | 0,273322 | 1,00000 | 0,118350 | 0,071282 | 0,252933 |
|  | Troponin I | Myoglobin | CKP-MB 1 | CKP-MB 2 | LDH 1 |
| RBC | 0,188032 |  | -0,082001 | -0,021656 | 0,001593 |
| lymphocytes | -0,034776 | 0,50000 | 0,063300 | -0,153597 | -0,061029 |
| Thrombocytes | 0,242812 |  | 0,164946 | 0,205310 | 0,095874 |
| ESR | -0,039481 | 0,50000 | 0,098663 | 0,013043 | 0,047350 |
| Ketones |  |  |  |  |  |
| Urine proteins | 0,045506 | 0,86603 | -0,044192 | -0,291265 | 0,137175 |
| leukocytes | 0,071426 | 0,50000 | 0,172589 | 0,219115 | 0,128269 |
| epithelial cells | 0,106241 | 0,50000 | 0,239036 | 0,329329 | 0,040671 |
| Troponin I | 1,000000 |  | 0,490390 | 0,265996 | 0,374820 |
| Myoglobin |  | 1,00000 |  |  |  |
| CKP-MB 1 | 0,490390 |  | 1,000000 | 0,386079 | 0,576087 |
| CKP-MB 2 | 0,265996 |  | 0,386079 | 1,000000 | 0,067851 |
| LDH 1 | 0,374820 |  | 0,576087 | 0,067851 | 1,000000 |
| LDH 2 | 0,319420 |  | 0,127235 | 0,437527 | 0,314912 |
| INR | 0,208372 |  | 0,057017 | 0,000653 | 0,141873 |
| Total bilirubin | 0,166543 |  | 0,189142 | 0,018849 | 0,112833 |
| ALT | 0,130182 |  | 0,152500 | 0,057059 | 0,200843 |
| AST | 0,170510 |  | 0,194561 | 0,053218 | 0,241813 |
| Total choles | 0,124187 | 0,50000 | -0,040978 | -0,006981 | 0,082099 |
| LDL | 0,195942 | 0,50000 | 0,061061 | 0,116077 | 0,097868 |
| TAG | 0,075288 | 0,50000 | -0,001916 | -0,046766 | 0,003054 |
| Na | 0,073038 | -1,00000 | -0,145310 | -0,202524 | 0,056657 |
| K | 0,111597 | 0,50000 | 0,007184 | -0,162865 | 0,162139 |
| Creatinine | 0,177284 | -1,00000 | 0,260587 | -0,008614 | 0,226690 |
| Ca | 0,002295 |  | -0,255494 | -0,448271 | -0,032190 |
| Uria | 0,077832 | 0,50000 | 0,050858 | -0,208204 | 0,114875 |
| Glucose | -0,027007 | 1,00000 | -0,062950 | -0,082192 | 0,112243 |
| APTT | 0,126145 |  | 0,309909 | 0,423780 | 0,087702 |
| Fibrinogen | -0,126763 |  | 0,019966 | -0,141352 | 0,038093 |
| PT |  |  |  |  |  |
|  | LDH 2 | INR | Total bilirubin | ALT | AST |
| Date of infarct | 0,123665 | 0,222657 | -0,070080 | -0,025792 | 0,033419 |
| Date of hosp. | 0,119836 | 0,210509 | -0,093753 | -0,033286 | 0,045471 |
| Date of dischargement | 0,113804 | 0,210587 | -0,091667 | -0,036998 | 0,045172 |
| Hospitalization days | -0,351822 | -0,122041 | 0,083439 | -0,149559 | -0,089148 |
| Age | -0,035860 | 0,088783 | 0,119390 | 0,006031 | 0,053983 |
| Body weight | -0,342356 | -0,087567 | 0,142107 | 0,357989 | 0,255829 |
| Height | 0,882919 | -0,115722 | 0,304978 | -0,136986 | 0,051121 |
| BMI | -0,357143 | 0,016422 | 0,013691 | 0,462577 | 0,191586 |
| HR | -0,065697 | -0,176321 | 0,091752 | 0,168135 | 0,058756 |
| SBP | -0,047346 | -0,009411 | -0,074120 | -0,014180 | -0,161133 |
| DBP | -0,023404 | -0,032399 | -0,152479 | -0,079591 | -0,161825 |
| Aorta diameter | -0,025929 | 0,012770 | 0,105600 | -0,081878 | 0,016592 |
| EF | -0,131816 | -0,041426 | -0,161993 | -0,068176 | -0,100612 |
| EDV | -0,114369 | 0,021536 | 0,154520 | 0,069499 | 0,069440 |
| LV | -0,150290 | -0,046075 | 0,007623 | 0,009479 | 0,068600 |
| LA | -0,053576 | 0,055728 | 0,182757 | 0,064334 | -0,049618 |
| RV | -0,082071 | 0,098168 | 0,195401 | 0,083873 | 0,001424 |
|  | LDH 2 | INR | Total bilirubin | ALT | AST |
| RA | 0,011156 | 0,162673 | 0,054035 | 0,022768 | 0,077034 |
| Hb | 0,254468 | 0,101077 | 0,215681 | 0,011558 | -0,009187 |
| leukocytes | 0,286870 | 0,187285 | -0,040948 | 0,067994 | 0,072187 |
| RBC | 0,126967 | 0,052727 | 0,089829 | 0,073779 | -0,080989 |
| lymphocytes | -0,211282 | -0,213453 | 0,074228 | -0,006331 | -0,038706 |
| Thrombocytes | 0,349153 | 0,200752 | -0,220295 | 0,035331 | -0,083374 |
| ESR | -0,210693 | -0,019873 | -0,048101 | 0,013972 | 0,000510 |
| Ketones |  |  |  |  |  |
| Urine proteins | 0,046534 | 0,145399 | 0,032055 | -0,037007 | 0,091987 |
| leukocytes | -0,120852 | -0,010482 | -0,140302 | -0,097826 | -0,132582 |
| epithelial cells | -0,084567 | -0,113416 | -0,083612 | -0,028856 | -0,041266 |
| Troponin I | 0,319420 | 0,208372 | 0,166543 | 0,130182 | 0,170510 |
| Myoglobin |  |  |  |  |  |
| CKP-MB 1 | 0,127235 | 0,057017 | 0,189142 | 0,152500 | 0,194561 |
| CKP-MB 2 | 0,437527 | 0,000653 | 0,018849 | 0,057059 | 0,053218 |
| LDH 1 | 0,314912 | 0,141873 | 0,112833 | 0,200843 | 0,241813 |
| LDH 2 | 1,000000 | 0,382124 | 0,002090 | 0,286460 | 0,447358 |
| INR | 0,382124 | 1,000000 | 0,107299 | -0,044216 | 0,049075 |
| Total bilirubin | 0,002090 | 0,107299 | 1,000000 | 0,175968 | 0,135787 |
| ALT | 0,286460 | -0,044216 | 0,175968 | 1,000000 | 0,572879 |
| AST | 0,447358 | 0,049075 | 0,135787 | 0,572879 | 1,000000 |
| Total choles | -0,049465 | -0,111364 | 0,135346 | 0,068525 | 0,067669 |
| LDL | -0,043774 | -0,011290 | 0,158629 | 0,020542 | 0,064199 |
| TAG | -0,033778 | -0,095456 | -0,039331 | 0,126804 | 0,040082 |
| Na | 0,126636 | 0,138965 | -0,085883 | -0,015813 | 0,130889 |
| K | -0,091874 | 0,175417 | 0,134458 | 0,035404 | 0,062157 |
| Creatinine | -0,170041 | 0,185665 | 0,069496 | -0,021837 | 0,002576 |
| Ca | -0,065550 | 0,229418 | -0,076928 | 0,098510 | -0,040195 |
| Uria | -0,156195 | 0,132454 | 0,048647 | 0,248900 | 0,155723 |
| Glucose | 0,089111 | 0,031761 | -0,009808 | 0,246838 | 0,156518 |
| APTT | 0,197419 | 0,167639 | 0,073983 | 0,084182 | -0,016993 |
| Fibrinogen | -0,089828 | -0,389201 | 0,105851 | -0,003601 | -0,044486 |
| PT |  |  |  |  |  |
|  | Total choles | LDL | TAG | Na | K |
| Date of infarct | -0,081606 | -0,120629 | 0,075539 | 0,29553 | 0,392714 |
| Date of hosp. | -0,083591 | -0,114472 | 0,041802 | 0,32023 | 0,347683 |
| Date of dischargement | -0,082453 | -0,110837 | 0,040081 | 0,32329 | 0,345315 |
| Hospitalization days | 0,135370 | 0,159497 | -0,018082 | -0,07405 | -0,119386 |
| Age | 0,189818 | 0,103719 | -0,089894 | 0,04271 | 0,096715 |
| Body weight | 0,032294 | 0,214156 | 0,190830 | 0,09980 | 0,262988 |
| Height | -0,138274 | -0,089237 | -0,346317 | -0,08695 | -0,486864 |
| BMI | 0,167920 | 0,287161 | 0,243163 | 0,14748 | 0,402687 |
| HR | 0,074696 | -0,019979 | -0,001737 | 0,01790 | 0,055393 |
| SBP | 0,130019 | 0,048523 | 0,159888 | 0,07313 | 0,000735 |
| DBP | 0,103124 | 0,051746 | 0,203202 | 0,02336 | -0,018963 |
| Aorta diameter | -0,272978 | -0,290000 | -0,064514 | 0,01220 | 0,099267 |
| EF | 0,001418 | -0,003403 | 0,135686 | 0,02820 | 0,047078 |
| EDV | -0,010848 | -0,023996 | -0,012581 | -0,06275 | 0,058684 |
|  | Total choles | LDL | TAG | Na | K |
| LV | -0,034502 | -0,070730 | 0,085614 | 0,04582 | -0,002292 |
| LA | -0,022993 | -0,008003 | 0,071785 | 0,14456 | 0,117294 |
| RV | -0,008355 | 0,006907 | 0,096231 | -0,05588 | 0,285462 |
| RA | -0,160652 | -0,091096 | -0,036389 | 0,18110 | 0,147145 |
| Hb | 0,066861 | 0,114681 | 0,157777 | 0,08106 | 0,218370 |
| leukocytes | -0,025760 | -0,024549 | 0,203652 | -0,06836 | 0,047010 |
| RBC | 0,023385 | 0,091089 | 0,172810 | -0,02729 | 0,187586 |
| lymphocytes | 0,111800 | 0,066237 | 0,157562 | 0,15747 | 0,193137 |
| Thrombocytes | -0,011878 | 0,051375 | -0,017568 | -0,15520 | -0,172012 |
| ESR | -0,128001 | -0,102900 | 0,038587 | -0,10175 | -0,033521 |
| Ketones |  |  |  |  |  |
| Urine proteins | -0,061160 | -0,057283 | 0,007271 | -0,07022 | -0,049361 |
| leukocytes | 0,106976 | 0,212155 | -0,001248 | -0,17767 | -0,202077 |
| epithelial cells | 0,121417 | 0,184569 | 0,020937 | -0,16269 | -0,275099 |
| Troponin I | 0,124187 | 0,195942 | 0,075288 | 0,07304 | 0,111597 |
| Myoglobin | 0,500000 | 0,500000 | 0,500000 | -1,00000 | 0,500000 |
| CKP-MB 1 | -0,040978 | 0,061061 | -0,001916 | -0,14531 | 0,007184 |
| CKP-MB 2 | -0,006981 | 0,116077 | -0,046766 | -0,20252 | -0,162865 |
| LDH 1 | 0,082099 | 0,097868 | 0,003054 | 0,05666 | 0,162139 |
| LDH 2 | -0,049465 | -0,043774 | -0,033778 | 0,12664 | -0,091874 |
| INR | -0,111364 | -0,011290 | -0,095456 | 0,13897 | 0,175417 |
| Total bilirubin | 0,135346 | 0,158629 | -0,039331 | -0,08588 | 0,134458 |
| ALT | 0,068525 | 0,020542 | 0,126804 | -0,01581 | 0,035404 |
| AST | 0,067669 | 0,064199 | 0,040082 | 0,13089 | 0,062157 |
| Total choles | 1,000000 | 0,830689 | 0,417038 | 0,09591 | -0,006258 |
| LDL | 0,830689 | 1,000000 | 0,319855 | 0,06534 | -0,075583 |
| TAG | 0,417038 | 0,319855 | 1,000000 | -0,00482 | 0,096150 |
| Na | 0,095906 | 0,065343 | -0,004822 | 1,00000 | 0,349223 |
| K | -0,006258 | -0,075583 | 0,096150 | 0,34922 | 1,000000 |
| Creatinine | 0,083018 | 0,087098 | -0,032333 | -0,01335 | -0,060077 |
| Ca | -0,108344 | -0,126906 | 0,066131 | 0,47663 | 0,575797 |
| Uria | 0,046621 | 0,055896 | 0,027175 | 0,09747 | 0,132856 |
| Glucose | 0,048354 | 0,026693 | 0,161067 | -0,03075 | 0,004122 |
| APTT | 0,110501 | 0,148558 | -0,068772 | -0,11314 | 0,049147 |
| Fibrinogen | 0,044268 | -0,029297 | 0,160949 | -0,09123 | 0,017443 |
| PT |  |  |  |  |  |
|  | Creatinine | Ca | Uria | Glucose | APTT |
| Date of infarct | -0,12362 | 0,752734 | 0,173533 | 0,198889 | -0,219685 |
| Date of hosp. | -0,10899 | 0,655580 | 0,172153 | 0,197077 | -0,213583 |
| Date of dischargement | -0,10773 | 0,652954 | 0,172156 | 0,199063 | -0,214560 |
| Hospitalization days | 0,11868 | -0,365114 | -0,032561 | -0,002611 | 0,142006 |
| Age | 0,19637 | -0,006410 | 0,189761 | 0,166185 | 0,049935 |
| Body weight | -0,00751 | 0,283974 | 0,121533 | 0,654202 | 0,003153 |
| Height | 0,03593 | -0,338311 | -0,404216 | -0,048985 | -0,204839 |
| BMI | -0,05175 | 0,422982 | 0,221784 | 0,497591 | 0,145463 |
| HR | -0,01873 | -0,077756 | 0,076878 | 0,077596 | 0,034608 |
| SBP | -0,11554 | -0,065510 | -0,020419 | -0,030442 | -0,055021 |
| DBP | -0,22954 | -0,085972 | -0,078703 | -0,022622 | -0,086966 |
| Aorta diameter | -0,13025 | -0,005432 | -0,032316 | 0,053932 | -0,118503 |
|  | Creatinine | Ca | Uria | Glucose | APTT |
| EF | -0,13633 | 0,020465 | 0,005515 | -0,146818 | -0,201638 |
| EDV | -0,09448 | -0,047146 | -0,078861 | 0,043452 | 0,173079 |
| LV | -0,06663 | -0,021513 | -0,037700 | 0,129861 | 0,040094 |
| LA | -0,02786 | 0,265162 | 0,189547 | 0,175194 | -0,129817 |
| RV | -0,20783 | 0,180772 | 0,194250 | 0,223505 | -0,000931 |
| RA | -0,10200 | 0,166642 | 0,101188 | 0,171877 | -0,139865 |
| Hb | -0,02468 | 0,194971 | -0,211940 | -0,008374 | 0,102761 |
| leukocytes | 0,06829 | 0,060749 | 0,037870 | 0,205235 | 0,104666 |
| RBC | -0,13715 | 0,269192 | -0,139107 | 0,090559 | 0,174735 |
| lymphocytes | -0,12491 | 0,198846 | -0,165130 | -0,125482 | 0,011001 |
| Thrombocytes | 0,18359 | -0,046922 | -0,004645 | -0,062960 | 0,316938 |
| ESR | -0,09753 | -0,090898 | 0,165846 | 0,021904 | -0,032073 |
| Ketones |  |  |  |  |  |
| Urine proteins | 0,03047 | 0,007566 | 0,190483 | 0,109950 | -0,184248 |
| leukocytes | 0,17726 | -0,376993 | 0,099496 | 0,067847 | 0,223590 |
| epithelial cells | 0,15818 | -0,433478 | 0,036004 | 0,160787 | 0,313909 |
| Troponin I | 0,17728 | 0,002295 | 0,077832 | -0,027007 | 0,126145 |
| Myoglobin | -1,00000 |  | 0,500000 | 1,000000 |  |
| CKP-MB 1 | 0,26059 | -0,255494 | 0,050858 | -0,062950 | 0,309909 |
| CKP-MB 2 | -0,00861 | -0,448271 | -0,208204 | -0,082192 | 0,423780 |
| LDH 1 | 0,22669 | -0,032190 | 0,114875 | 0,112243 | 0,087702 |
| LDH 2 | -0,17004 | -0,065550 | -0,156195 | 0,089111 | 0,197419 |
| INR | 0,18567 | 0,229418 | 0,132454 | 0,031761 | 0,167639 |
| Total bilirubin | 0,06950 | -0,076928 | 0,048647 | -0,009808 | 0,073983 |
| ALT | -0,02184 | 0,098510 | 0,248900 | 0,246838 | 0,084182 |
| AST | 0,00258 | -0,040195 | 0,155723 | 0,156518 | -0,016993 |
| Total choles | 0,08302 | -0,108344 | 0,046621 | 0,048354 | 0,110501 |
| LDL | 0,08710 | -0,126906 | 0,055896 | 0,026693 | 0,148558 |
| TAG | -0,03233 | 0,066131 | 0,027175 | 0,161067 | -0,068772 |
| Na | -0,01335 | 0,476633 | 0,097467 | -0,030751 | -0,113135 |
| K | -0,06008 | 0,575797 | 0,132856 | 0,004122 | 0,049147 |
| Creatinine | 1,00000 | -0,021655 | 0,477372 | 0,009566 | 0,130883 |
| Ca | -0,02165 | 1,000000 | 0,226440 | 0,114659 | -0,213578 |
| Uria | 0,47737 | 0,226440 | 1,000000 | 0,223992 | -0,038725 |
| Glucose | 0,00957 | 0,114659 | 0,223992 | 1,000000 | -0,091630 |
| APTT | 0,13088 | -0,213578 | -0,038725 | -0,091630 | 1,000000 |
| Fibrinogen | -0,03245 | -0,074313 | 0,054461 | 0,016700 | -0,101748 |
| PT |  |  |  |  |  |

| Variable | Pearson Rank Order Correlations (Acute MI in Total Post MI 5)<br>MD pairwise deleted<br>Marked correlations are significant at p <,05000 |  |
| --- | --- | --- |
|  | Fibrinogen | PT |
| Date of infarct | 0,070849 |  |
| Date of hosp. | 0,061743 |  |
| Date of dischargement | 0,060104 |  |
| Hospitalization days | 0,014194 |  |
| Age | -0,101096 |  |
| Body weight | -0,200220 |  |
| Height | -0,219706 |  |

| Variable | Pearson Rank Order<br>Correlations (Acute MI in<br>Total Post MI 5)<br>MD pairwise deleted<br>Marked correlations are<br>significant at $p < ,05000$ | |
| --- | --- | --- |
|  | Fibrinogen | PT |
| BMI | -0,051268 |  |
| HR | -0,068684 |  |
| SBP | 0,014181 |  |
| DBP | -0,081389 |  |
| Aorta diameter | -0,066697 |  |
| EF | 0,067573 |  |
| EDV | -0,119314 |  |
| LV | -0,116229 |  |
| LA | 0,006765 |  |
| RV | -0,007770 |  |
| RA | -0,072335 |  |
| Hb | -0,051919 |  |
| leukocytes | -0,093511 |  |
| RBC | 0,020243 |  |
| lymphocytes | 0,044111 |  |
| Thrombocytes | -0,224921 |  |
| ESR | -0,019113 |  |
| Ketones |  |  |
| Urine proteins | 0,032734 |  |
| leukocytes | 0,074379 |  |
| epithelial cells | 0,100294 |  |
| Troponin I | -0,126763 |  |
| Myoglobin |  |  |
| CKP-MB 1 | 0,019966 |  |
| CKP-MB 2 | -0,141352 |  |
| LDH 1 | 0,038093 |  |
| LDH 2 | -0,089828 |  |
| INR | -0,389201 |  |
| Total bilirubin | 0,105851 |  |
| ALT | -0,003601 |  |
| AST | -0,044486 |  |
| Total choles | 0,044268 |  |
| LDL | -0,029297 |  |
| TAG | 0,160949 |  |
| Na | -0,091235 |  |
| K | 0,017443 |  |
| Creatinine | -0,032451 |  |
| Ca | -0,074313 |  |
| Uria | 0,054461 |  |
| Glucose | 0,016700 |  |
| APTT | -0,101748 |  |
| Fibrinogen | 1,000000 |  |
| PT |  | 1,000000 |

|  |  |
| --- | --- |
| Variable | Correlations<br>(Acute MI in<br>Total Post MI 5)<br>Marked<br>correlations are<br>significant at p<br>< ,05000<br>N=30 (Case<br>wise deletion of<br>missing data) |
|  | Body weight |
| BMI | 0,895885 |

